# The rise and spread of the SARS-CoV-2 AY.122 lineage in Russia

**DOI:** 10.1101/2021.12.02.21267168

**Authors:** Galya V. Klink, Ksenia Safina, Elena Nabieva, Nikita Shvyrev, Sofya Garushyants, Evgeniia Alekseeva, Andrey B. Komissarov, Daria M. Danilenko, Andrei A. Pochtovyi, Elizaveta V. Divisenko, Lyudmila A. Vasilchenko, Elena V. Shidlovskaya, Nadezhda A. Kuznetsova, The Coronavirus Russian Genetics Initiative (CoRGI) Consortium, Andrei E. Samoilov, Alexey D. Neverov, Anfisa V. Popova, Gennady G. Fedonin, The CRIE Consortium, Vasiliy G. Akimkin, Dmitry Lioznov, Vladimir A. Gushchin, Vladimir Shchur, Georgii A. Bazykin

## Abstract

**Background:** Delta has outcompeted most preexisting variants of SARS-CoV-2, becoming the globally predominant lineage by mid-2021. Its subsequent evolution has led to emergence of multiple sublineages, many of which are well-mixed between countries.

**Aim:** Here, we aim to study the emergence and spread of the Delta lineage in Russia.

**Methods:** We use a phylogeographic approach to infer imports of Delta sublineages into Russia, and phylodynamic models to assess the rate of their spread.

**Results:** We show that nearly the entire Delta epidemic in Russia has probably descended from a single import event despite genetic evidence of multiple Delta imports. Indeed, over 90% of Delta samples in Russia are characterized by the nsp2:K81N+ORF7a:P45L pair of mutations which is rare outside Russia, putting them in the AY.122 sublineage. The AY.122 lineage was frequent in Russia among Delta samples from the start, and has not increased in frequency in other countries where it has been observed, suggesting that its high prevalence in Russia has probably resulted from a random founder effect.

**Conclusion:** The apartness of the genetic composition of the Delta epidemic in Russia makes Russia somewhat unusual, although not exceptional, among other countries.

## Introduction

In a pandemic, the global spread of viral lineages is defined by a multitude of factors including the intrinsic properties of the virus, properties of host populations, social factors and chance. Distinguishing between these factors remains challenging; in particular, it is difficult to spot the lineages with increased fitness against the background of random frequency fluctuations. Since the start of the SARS-CoV2 pandemic, several lineages of concern have appeared and replaced preexisting lineages in different countries [1]. While some of these variants are certainly characterized by changed fitness due to changes in transmissibility and/or immune avoidance [2], much of the geographical difference and dynamics of SARS-CoV-2 lineages is due to epidemiological factors that are not caused by differences in variant fitness [3–5].

The Delta (B.1.617.2 + AY.*) variant of SARS-CoV-2 that was first detected in India in late 2020 [6] is, as of November 2021, the prevalent lineage in most countries including Russia [7]. It was shown to be more infectious but also to cause higher mortality than earlier variants of concern [8,9]. The fast spread of Delta may be associated with its reduced sensitivity to neutralization by antibodies [10] as well as increased efficiency of fusion with human cells [11]. Delta has spread rapidly in Russia, increasing in frequency from 1% in April to over 90% in June [12,13].

The phylogeny of Delta is more structured than that of other variants of concern, and its characteristic mutations have accumulated gradually [14]. While Delta clearly has increased fitness compared to ancestral strains, whether its sublineages change its properties further is less clear [15–17]. Still, adaptive evolution of SARS-CoV-2 in the human genome continues [18], highlighting the need for surveillance of novel variants.

Thanks to extensive efforts of many countries in sampling and sequencing SARS-CoV-2 genomes from patients, it is possible to track the spread of different viral variants across the world. Here, we analyze the emergence and spread of the Delta variant in Russia between April-October 2021 and compare it to other countries. We show that the majority of Russian samples carry the same set of mutations, strongly suggesting that they have descended from a single import event.

## Materials and Methods

### Data

We downloaded a masked alignment of 4,452,413 SARS-CoV2 sequences from GISAID on October 21, 2021 together with accompanying metadata. We retained sequences characterized as follows: “Variant” = “VOC Delta GK/478K.V1 (B.1.617.2+AY.x) first detected in India”, “Host” = “Human”, “Is complete?” = “True” and “Is high coverage?” = “True”. 1,439 Russian and 1,428,049 non-Russian samples were retained for analysis.

### UShER phylogenetic tree

We downloaded the public UShER mutation-annotated tree together with metadata on September 21, 2021 from the UCSC browser (http://hgdownload.soe.ucsc.edu/goldenPath/wuhCor1/UShER_SARS-CoV-2/). To avoid duplicate entries, we removed the Russian sequences present in the UShER tree, and then added the Russian GISAID sequences to the tree using UShER [19]. Branch lengths were corrected using mutation paths obtained by matUtils [20].

### Maximum likelihood phylogenetic trees

We created ten subsampled datasets by combining ten random subsets of 50,000 non-Russian Delta sequences with all Russian Delta sequences and with the hCoV-19/Australia/VIC18574/2021 sample from the B.1.617.1 lineage as an outgroup. For each dataset, we built a maximum likelihood phylogenetic tree using the FastTreeDbl algorithm of FastTree 2.1.11 [21] with the GTR substitution model and gamma model for heterogeneity of evolutionary rates across sites. We rooted the trees, and collapsed branches with less than one mutation (i.e. with length below 0.00003 mutations per site).

### Phylogenetic inference of imports

Imports into Russia were inferred from the phylogenetic distribution of sequences as follows (Figure S1). Samples (tree tips) were marked as Russian (*R*) or non-Russian (*O*) by place of collection. All internal nodes were numbered in order along each lineage from root to tip. Moving from the nodes with the highest numbers towards the lowest (root), each node N was labelled according to the labels of its immediate descendants (tips or internal nodes) as follows: (i) if more than one descendant was labelled *R*, N was labelled *R*; (ii) if no descendants were labelled *R*, N was not labelled; (iii) if exactly one descendant was labelled *R*, the branch leading to this descendant was marked as an import, and N was not labelled. As many of the phylogenetic branches are very short and often comprise just one mutation, we found that nucleotide miscalling can result in phylogenetic misplacement of samples and therefore erroneous inference of imports. To focus on the most robust imports, for nested import events, only the deepest import was retained. Imports into other countries were identified analogously. The python script for finding imports is available on GitHub: https://github.com/GalkaKlink/Delta-lineage-in-Russia

### Estimation of the logistic growth rates

Logistic growth rates of the Delta lineage were estimated with the nls() function of the R language [22]. For this, Delta frequencies among Russian samples were averaged across 15-days sliding windows (spanning the 7 days before the current date, the current date, and the 7 days after the current date), and windows with fewer than 20 samples were filtered out. Confidence intervals for estimated model parameters were calculated with confint2() function from nlstools package [23].

### Estimation of the effective reproduction number

We used the skyline birth-death model (BDSKY) [24] with continuous sampling, or ψ-sampling, implemented in BEAST2 [25] to infer the dynamics of the effective reproduction number R_e_. We focused on the monophyletic clade corresponding to the major Delta import. To tackle sampling heterogeneity, we filtered the major clade in two steps. First, we limited our analysis to the samples collected in Moscow. Second, we subsampled overrepresented dates (see Figure S2) in the Moscow dataset, because it violates the ψ-sampling model assumptions. Overrepresented dates most likely correspond to additional day-specific sampling events in contrast with continuous routine sampling. The additional batches generated at these dates should be described by ρ-sampling (sampling with a certain probability at a specific time). To remove biases from this overrepresentation, we downsampled the dataset so that it would fit with continuous ψ-sampling using the following procedure. For each date with at least ten samples, we calculated the mean number N of samples in a two week interval (one week before and one week after the date). Then we randomly kept k * N samples for this date with k=1 for the baseline analysis (see Figure S2), and additionally with k=0.5 and 2 to check the robustness of our procedure. Our results were not sensitive to k (Figure S6). Analyses were run for 100 million MCMC steps; convergence was assessed in Tracer [26]. We used the skylinetools package (https://github.com/laduplessis/skylinetools) to set monthly time points for the reproduction number and sampling proportion. All priors were kept default except for those provided in Table S1.

### Estimation of relatedness

To measure the relatedness of samples from the same country, we calculated the mean phylogenetic distance (distance along the phylogenetic tree, 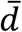) between 100 random pairs of samples from this country and compared it with the distribution of phylogenetic distances between 1000 random pairs of samples from any country. We then calculated the number of standard deviations (standard score) between 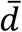 and the mean of this distribution; negative standard score corresponds to increased relatedness of samples from the same country, and positive score, to decreased relatedness. Scripts for phylogenetic clustering estimation are available on GitHub: https://github.com/GalkaKlink/Delta-lineage-in-Russia

### Visualization

The following R packages were used for visualization: tidyverse [27], ggrepel [28], egg [29], stringr [30] and Hmisc [31]. Phylogenetic tree was visualized using the ete3 framework [32].

## Results

### Delta has spread in Russia rapidly in spring 2021

Among the 4,639 high-quality Russian samples that were available in GISAID on October 21, 2021, 1,439 are Delta samples, i.e., belong to pango lineage B.1.617.2 or derived lineages (AY.*). The earliest high-quality Delta sample was collected on April 7, 2021 in Moscow; two lower-quality Delta samples date to February 28 and March 26, 2021. Since April, the frequency of Delta among the Russian samples has been growing, reaching 98% by early July 2021, with the estimated daily logistic growth rate of 9.74% (95% C.I.: 9.28%-10.2%). This growth rate is comparable with that observed in other countries [33,34]. The timing of this growth was similar between Russia’s regions (Fig. 1).

**Figure 1.**
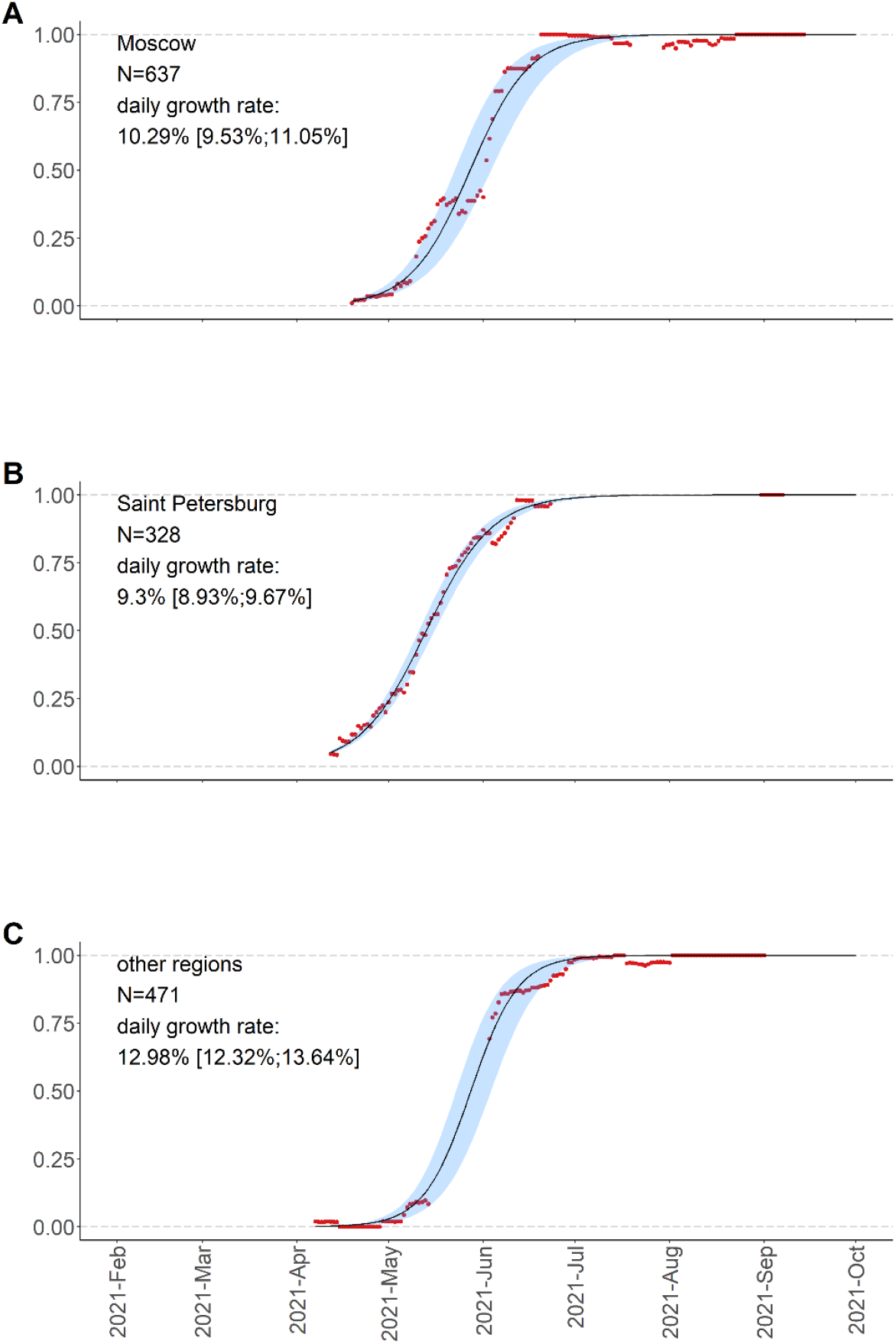
Frequencies of Delta variants (B.1.617.2+AY.*) in Russia measured for 15-day sliding windows of 7 days around each day, and logistic growth estimates with 95% confidence intervals.

### Most Russian Delta samples are characterized by the nsp2:K81N+ORF7a:P45L combination of mutations

The vast majority of Russian Delta samples shared the same combination of mutations (Fig. 2, Fig. S3). In addition to the mutations characteristic of Delta [7], 92.4% of the Delta samples carried the nsp2:K81N (ORF1a:K261N) mutation, and 91.8% carried the ORF7a:P45L mutation. The presence of the nsp2:K81N mutation puts these 92.4% of Russian Delta samples in the recently designated AY.122 pango lineage. The nsp2:K81N+ORF7a:P45L combination is rare among GISAID Delta samples worldwide (2.3%); outside Russia, its frequency is the highest in Moldova (100%; 9 out of 9 samples), followed by Ecuador (86%; 76 out of 89 samples), Kazakhstan (76%; 32 out of 42 samples) and Latvia (73%; 52 out of 71 samples).

**Figure 2.**
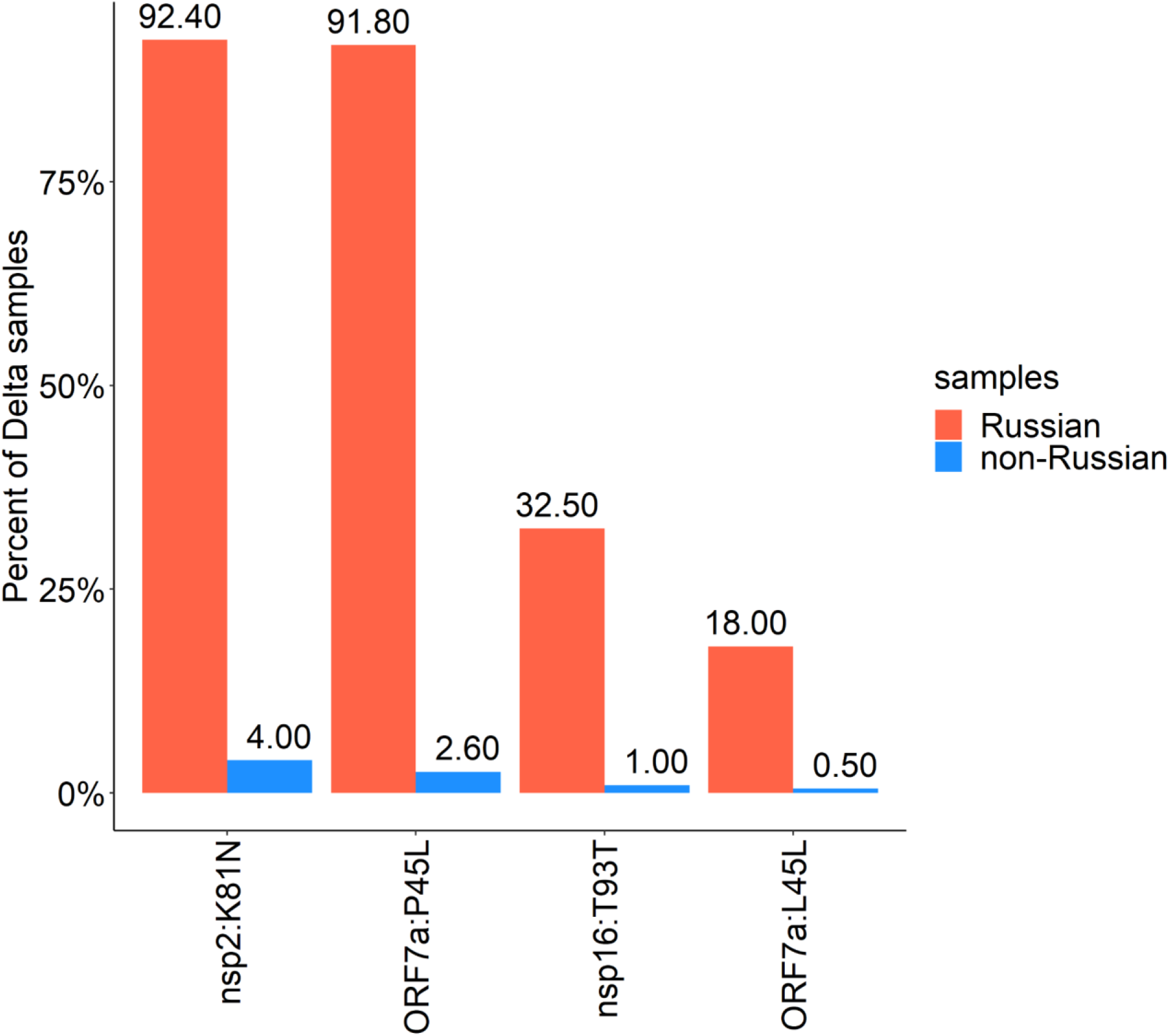
Mutations in the Delta lineage observed in >5% of Russian Delta samples. The following mutations that characterize the major sublineage of B.1.617.2 (“21J” in Nextstrain nomenclature) and occur in >85% of Delta samples both in Russia and globally are not shown: RdRp:G671S, exonuclease:A394V, nsp6:T77A, nsp3:A488S, nsp3:P1228L, nsp6:V120V, ORF7b:T40I, nsp3:P1469S, N:G215C, nsp4:D144D, nsp4:V167L, and nsp4:T492I.

Outside Russia, the nsp2:K81N and ORF7a:P45L mutations are not strongly linked, and many samples carry the first but not the second (Fig. 2, Fig.S3). The ORF7a:P45L mutation has been gained and lost repeatedly according to the global UShER tree. Notably, it is located within one of the ARTIC primers (nCoV-2019_90_RIGHT) binding site, suggesting that the nucleotide at this position may be frequently miscalled. However, in the Russian dataset, we find that the linkage between nsp2:K81N and ORF7a:P45L is nearly perfect, and these mutations cooccur in nearly all samples (Fig. 2, Fig.S3).

The earliest nsp2:K81N+ORF7a:P45L sample in Russia dates to April 19th, and it was one of the first Delta samples obtained in Russia. The frequency of the nsp2:K81N+ORF7a:P45L combination has been steadily high between April and October, and it remained the dominant clade throughout this period (Fig. 3A).

**Figure 3.**
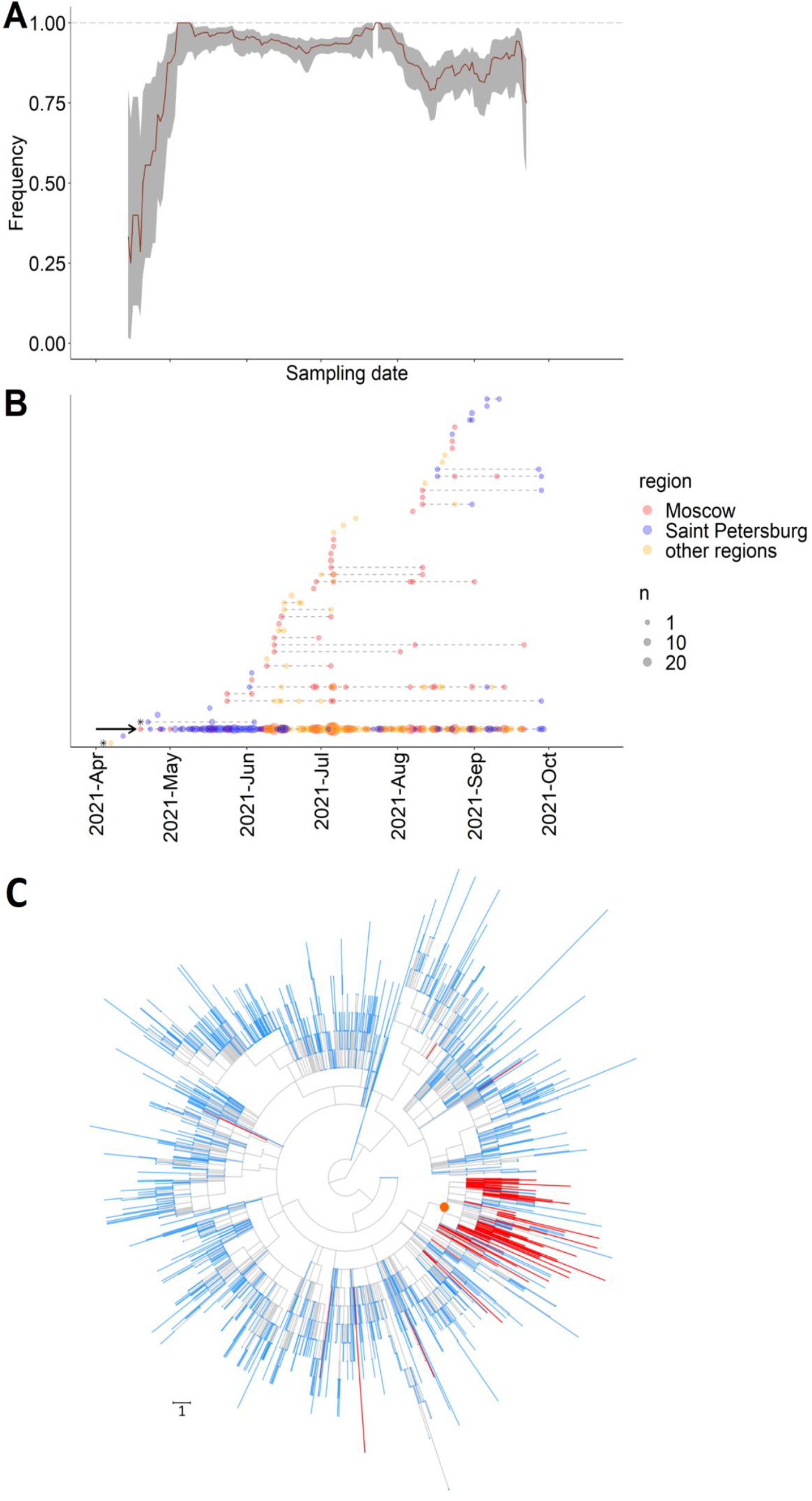
Dynamics of Delta sublineages in Russia. A) The fraction of the nsp2:K81N+ORF7a:P45L combination among all Delta samples from Russia in 15-day sliding window. The confidence band is the 95% binomial confidence interval. B) Timeline for imports of Delta subclades into Russia. Each horizontal line represents a Russian subclade descendant from one import, ordered by the date of the earliest sample. Circles represent samples taken on a particular date; circle size reflects the number of samples; circle color indicates the region of sampling. The AY.122+ORF7a:P45L sublineage is marked by an arrow. The two imports with known travel history are shown as asterisks. C) AY.122+ORF7a:P45L sublineage on the UShER tree of Delta. For visualization purposes, 90% of Russian and 99% of non-russian tips were pruned randomly. An internal node that was defined as the main import and which defines the AY.122+ORF7a:P45L sublineage is marked by an orange circle; Russian samples are colored in red; non-Russian samples are colored in blue; internal branches are colored in grey. Branch lengths are measured in the number of mutations.

Soon after its first detection, the nsp2:K81N+ORF7a:P45L combination has become prevalent throughout Russia (Fig. S4, S5). It was detected in all 41 Russian regions where Delta samples were collected. In the 26 regions with more than five samples of Delta, between 62% and 100% of samples carried the nsp2:K81N+ORF7a:P45L combination (Table S2).

### Just one Delta lineage was successful in Russia although many were imported

To understand how Delta variants were imported into Russia, we used a phylogeographic analysis. Using UShER [19], we constructed a global phylogeny of SARS-CoV-2 Delta samples including all 1,439 Delta specimens from Russia obtained between April 7 - September 29, 2021. In a maximum parsimony-based approach, we then identified import events as branches in the phylogenetic tree leading to the clades consisting of Russian samples such that their sister clades are non-Russian. For phylogenetically nested imports, only the deepest events were considered (Fig. S1; see Methods). Our procedure for detection of imports is conservative in that it does not allow repeated imports along the same phylogenetic lineage. It generally yields fewer imports than an alternative approach using Treetime [35], but the imports detected using maximum parsimony are nearly always also supported by Treetime [36]. The imports inferred under this definition matched well the clusters of Russian sequences observed in phylogenies.

Using this procedure, we detected 50 imports of the Delta lineage. 24 of these imports are represented by a single sequenced Russian sample each, while each of the remaining 26 is represented by multiple Russian samples descending from them. For two early events, the first samples have known travel histories (Fig. 3B). One of them was obtained on April 7th from a person who travelled to the UAE and Turkey, and this was the earliest high-quality Russian sample of the Delta lineage. The other was obtained on April 22nd from a person who travelled to India.

Strikingly, 91.2% of all samples descended from just a single import (hereafter referred to as the “main import”) characterised by the nsp2:K81N+ORF7a:P45L combination of mutations (Fig. 3B,C). Therefore, phylogenetic inference indicates that this pair of mutations has a common origin in Russia. The first sample from the main import was collected on April 19, 2021 in Moscow. The main import was among the earliest imports of Delta in Russia (Fig. 3B).

Phylogenetic inference of imports is sensitive to details of sampling and phylogenetic reconstruction. To estimate the robustness of our estimates, we validated them using an alternative phylogenetic approach. For this, we used the 1,428,049 non-Russian Delta sequences that were available in GISAID on October 21, 2021 after quality filtering (see Methods). We generated ten subsets of 50,000 random non-Russian samples with all 1,439 filtered Russian samples added, and reconstructed the maximum likelihood (ML) phylogenetic trees for each such subsample. The inferred number of imports differed between replicates, mainly due to low robustness of the smaller imports. Nonetheless, in each of the phylogenetic replicates, over 90% of Delta samples were inferred to be descendants of a single import event (Table 1), similarly to the results obtained with the UShER tree.

**Table 1.** Imports into Russia estimated on ten independent ML phylogenetic trees

| ML-tree | number of imports | Russian samples in biggest import | % Russian samples in largest import | first import (earliest sample) | last import (earliest sample) | largest import (earliest sample) |
| --- | --- | --- | --- | --- | --- | --- |
| 1 | 36 | 1315 | 0.92 | 2021-04-07 | 2021-09-06 | 2021-04-19 |
| 2 | 7 | 1419 | 0.99 | 2021-04-07 | 2021-09-06 | 2021-04-07 |
| 3 | 11 | 1386 | 0.97 | 2021-04-07 | 2021-07-15 | 2021-04-12 |
| 4 | 13 | 1386 | 0.97 | 2021-04-07 | 2021-07-15 | 2021-04-12 |
| 5 | 10 | 1386 | 0.97 | 2021-04-07 | 2021-07-15 | 2021-04-12 |
| 6 | 9 | 1396 | 0.98 | 2021-04-07 | 2021-08-11 | 2021-04-12 |
| 7 | 5 | 1408 | 0.99 | 2021-04-07 | 2021-08-17 | 2021-04-07 |
| 8 | 31 | 1319 | 0.92 | 2021-04-07 | 2021-09-06 | 2021-04-19 |
| 9 | 5 | 1379 | 0.97 | 2021-04-07 | 2021-08-11 | 2021-04-12 |
| 10 | 6 | 1389 | 0.97 | 2021-04-07 | 2021-08-11 | 2021-04-12 |

### Phylodynamics of the main import clade

To infer the rate of spread of the largest introduced Delta sublineage, we performed its phylodynamic analysis using BEAST2 [25]. COVID-19 has hit Russia’s regions differently and non-synchronously; for example, the timing of epidemic waves has differed among regions (https://xn--80aesfpebagmfblc0a.xn--p1ai/information/). To minimize any effects of geographic structure, for this analysis, we focused on a single region. We considered the 333 samples collected in Moscow, which is the best-sampled of all Russia’s regions.

The phylodynamic estimate of R_e_ of the main import clade was 2.16 (95% CI [1.81-2.52]) in May, and 1.27 (95% CI [1.09-1.44]) in June. In July, it dropped to 0.62 (95% CI [0.44-0.81]), and rose again to 1.00 (95% CI [0.79-1.20]) in August and 1.28 (95% CI [0.63-1.95]) in September, the last month covered by our genetic analysis (Fig. 4).

**Figure 4.**
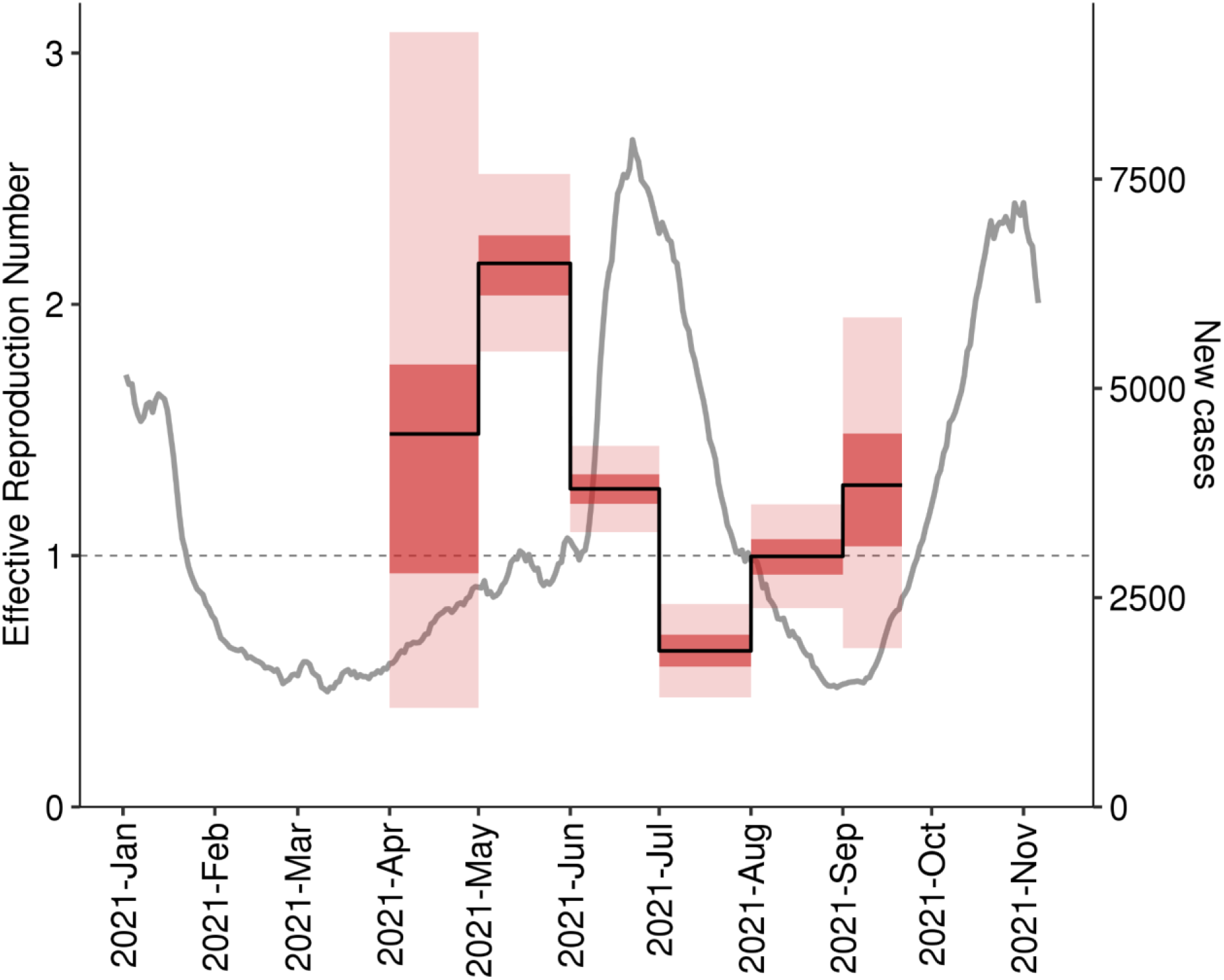
The estimated trajectory of the effective reproduction number R_e_ for the main import Delta clade in Moscow. The gray line shows the 7-day rolling average of the daily number of new cases (independent of genotype) in Moscow.

Overall, this dynamic was consistent with epidemiological data, with increases in R_e_ preceding rises in case counts (Fig. 4). Notably, the case counts before June include a large proportion of non-Delta cases; the reduction in number of non-Delta cases may partially explain why the rise in cases in May was slower than that predicted by the R_e_. Nevertheless, the high R_e_ in May and June is consistent with the summer wave which peaked on June 25, and the low R_e_ in July is consistent with the decline in case counts at that time (Fig. 4). This data confirms that the main import clade (AY.122) is responsible for the summer epidemic wave, and most probably for the ongoing autumn wave.

### The success of the AY.122+ORF7a:P45L combination is probably not due to increased fitness

To explain the success of the nsp2:K81N+ORF7a:P45L combination in Russia, we hypothesized that it could arise from fitness advantage conferred by these two mutations.

The identity of these mutations does not lend strong support to this hypothesis. nsp2 is a rapidly evolving non-structural protein which was found to be localized to endosomes and viral replication-transcription complexes. Based on structural analysis and affinity purification mass spectrometry, it is thought to interact with multiple host proteins and mitochondrial RNA, and its suggested functions are engagement of mitochondria to viral replication sites and modulation of cellular endosomal pathway [37]. No signs of either positive or negative selection were found at site 81 of nsp2 (https://observablehq.com/@spond/evolutionary-annotation-of-sars-cov-2-covid-19-genomes-enab?collection=@spond/sars-cov-2) using FEL and MEME algorithms of HyPhy [38].

ORF7a has been shown to suppress BST2 protein that restricts the egress of viral particles from the cell [39]. It was also shown to bind to CD14+ monocytes, which reduces their antigen representation capacity and triggers the production of proinflammatory cytokines [40]. Nonsynonymous mutations in ORF7a contribute to SARS-CoV-2 clade success [18]. C-terminal truncations of ORF7a are frequent and were shown to affect viral replication [41]. Nevertheless, a lineage characterized by a frameshifting deletion in ORF7a has spread rapidly in Australia [42]. Site ORF7a:45 experiences episodic diversifying selection (according to MEME algorithm of HyPhy) and increase of non-reference amino acid in frequency according to (https://observablehq.com/@spond/evolutionary-annotation-of-sars-cov-2-covid-19-genomes-enab?collection=@spond/sars-cov-2). It has also been predicted to be included in the B-cell epitope [43].

Moreover, the dynamics of the nsp2:K81N + ORF7a:P45L combination outside Russia also doesn’t support its increased fitness. To show this, we estimated the logistic growth rates of this combination in those countries where it has been frequent (with >15 days with samples carrying this combination both before and after July 1). While this lineage has been growing in most countries before July 1 (Fig. S7), this growth was due to the weakness of competition from non-Delta variants; no systematic growth compared with other Delta lineages was observed (Fig. S8). The lack of a systematic fitness advantage of this lineage across the globe suggests that the selection that favors this variant, if it exists, is weak.

### The genetic homogeneity of Delta in Russia is unusual among other countries

To compare the genetic uniformity of Delta samples observed in Russia to that in other countries, we used the same procedure to obtain a list of import events for each country with more than 50 Delta sequences in each ML tree. For each country, we then calculated (i) the fraction of Delta samples descendant from the largest import into this country, and (ii) the extent of relatedness of Delta samples from this country, compared to randomly chosen Delta samples (see Methods).

The contribution of the largest import was higher in Russia than in most other countries (Fig. 5A). Moreover, while samples from most countries were scattered across the phylogenetic tree, with multiple imports contributing substantially to the local epidemics, Russian samples were unusually related (Fig. 5B). Both these observations also held for the UShER trees that were based on smaller open datasets (Fig. S9).

**Figure 5.**
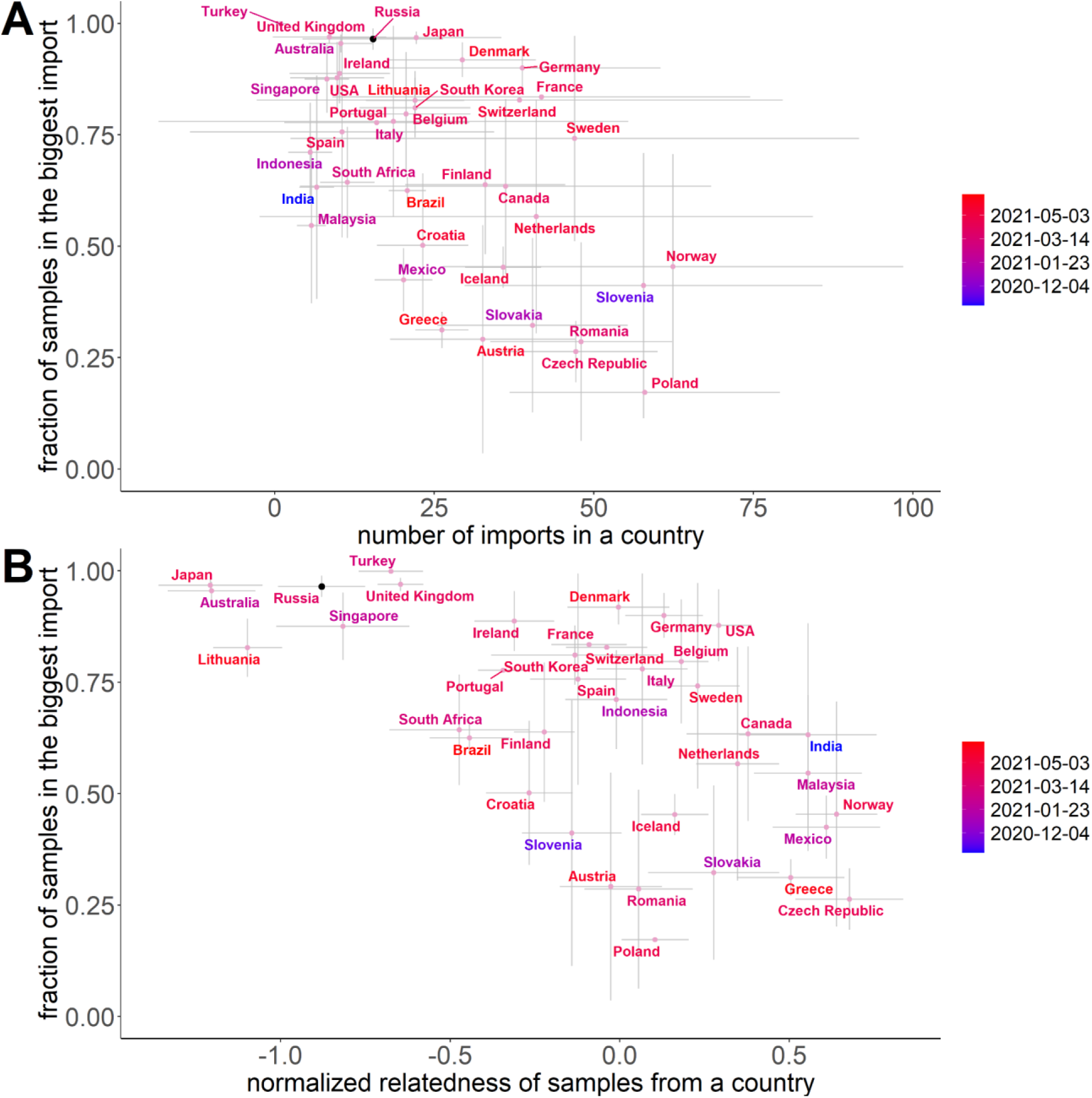
Fraction of Delta samples in the largest import and relatedness of Delta samples, for countries with at least 50 Delta samples in each of the 10 ML trials (Table S3). In (B), the horizontal axis indicates the normalized relatedness of samples from the same country, compared with randomly picked samples; lower values correspond to increased relatedness (see Methods). Dots correspond to the mean (centroid) across the 10 ML trees for 50,000 non-Russian samples with added Russian sequences, with standard deviations shown as error bars. Colors indicate the date when the Delta lineage reached 1% frequency in this country.

## Discussion

Previously, we and others have shown that transmission of pre-Delta SARS-CoV-2 variants across Russia’s border was rapid [44,45]. Indeed, the COVID-19 epidemic was started in Russia by a large number of near-simultaneous imports of distinct variants in early spring 2020, and many of these imports resulted in sizable Russian transmission lineages with no single lineage dominating [45]. In subsequent months, imports have continued despite border closure, resulting in thousands of Russian transmission lineages [36].

By contrast, here we show that the vast majority of Delta SARS-CoV-2 variants that have spread in Russia were genetically similar, carrying the derived nsp2:K81N and ORF7a:P45L changes that are rare outside Russia.

Our ability to distinguish between viral variants resulting from specific imports is limited by the resolution provided by genomic sequences. It is impossible to distinguish between repeated imports of genetically similar or identical variants, and this could lead us to undercount imports. However, the finding of the biased composition of the Russian Delta epidemic does not depend on this concern. At the time of import of the major Delta lineage into Russia, the global diversity of Delta variants was already high, and we would have been able to identify distinct Delta variants. Indeed, the nsp2:K81N+ORF7a:P45L combination occurs in 68 out of 80 (85%) of Russian samples obtained in April, but just in 34 out of 6658 (0.5%) of non-Russian samples obtained at that time.

What can account for this uniformity? There are several options. Conceivably, these mutations could increase viral fitness. Both mutations characterizing the main import clade, nsp2:K81N and ORF7a:P45L, are nonsynonymous, making this possibility realistic. However, neither of the two mutations is an obvious candidate for adaptiveness. There is also no evidence that the nsp2:K81N+ORF7a:P45L combination is characterized by an increased rate of spread compared to other Delta variants. While AY.122 has spread rapidly throughout Russia, and at least in Moscow this spread has been driven by a high R_e_>1 (Fig. 4), this rapid spread was against the background of non-delta variants (Fig. 1). The frequency of AY.122 among delta variants in Russia has been high almost from the start, and its early increase in frequency among Delta samples (Fig. 3A) happened while Delta cases were still low, indicating that it could be random. While it has later been established in numerous other countries in late spring and summer, its frequency has remained modest and not increased monotonically outside Russia.

Therefore, the high prevalence of the AY.122+ORF7a:P45L lineage in Russia is probably due to chance. As dispersal of SARS-CoV-2 within countries is more rapid than trans-border transmission, the role of chance in the spread of selectively equivalent variants is high. Indeed, some of the lineages have previously risen in frequency in Russia (e.g. B.1.1.317, [46]) and elsewhere (e.g. “European lineage” EU1, [47]) before declining, indicating that the changes defining them did not confer substantial fitness advantage. Although imports of Delta variants into Russia were multiple, even closely dated imports differed strikingly by their success (Fig. 3B).

Several factors could contribute to the biased composition of the Delta epidemic in Russia compared to most other countries. First, it could result from a geographic bias in the origin of imports. The earliest samples carrying the nsp2:K81N+ORF7a:P45L combination are sporadic, and were often deposited months later than collected, suggesting that they could be misdated in GISAID (Table S4). However, since mid-April, this combination started to appear nearly simultaneously among Delta samples from multiple countries. This suggests that it had originated earlier in a poorly sampled location (Table S4). In the second quarter of 2021, the top ten countries with the strongest passenger traffic with Russia were Abkhazia, Ukraine, Turkey, Kazakhstan, UAE, Cyprus, Armenia, Finland, South Ossetia, and Egypt (https://fedstat.ru/indicator/38480). Most of these countries sequence little. Nevertheless, nsp2:K81N+ORF7a:P45L has been observed in four of them (Table S4).

Second, the size of the trans-border transmission bottleneck could be affected by the measures aimed at limiting passenger traffic. Indeed, the countries with the largest contribution of a single import event to Delta cases (those in top left in Fig. 5B) include Japan, Australia and Singapore, some of the countries with the most stringent border policies in place. In Russia, however, the situation was very different. International traffic through Russian airports was higher in Spring 2021 than in most months of 2020, and rose to pre-pandemic levels by late 2021 (Table S5). Novel pre-Delta transmission lineages have been emerging in Russia right up to the moment when Delta appeared (Fig. S10). Delta has also been introduced into Russia repeatedly (Fig. 3B), indicating that the homogeneity of the epidemic results from a high variance of reproductive success of imports rather than from their low numbers.

Third, the success of the AY.122+ORF7a:P45L lineage in Russia could arise from an early superspreading event. Generally, superspreading events have been crucial for SARS-CoV2 spread [4]. However, no such event was reported. The AY.122+ORF7a:P45L variant started to spread near-simultaneously in Moscow, Saint Petersburg and the remainder of Russia (Table S2), suggesting that if true, this event took place before April 19 in a poorly sampled location within or outside Russia.

Independent of its exact mechanism, the high prevalence of just a single Delta variant in Russia highlights the high role of chance in the local spread of pathogenic lineages. This is in line with the high variance in levels of genetic differentiation (Fst) between countries early in the COVID-19 pandemic, suggesting that outbreaks in most countries could have been started by just a handful of travellers [48]. It takes few imports to start an epidemic.

## Supporting information

List of CORGI members

List of CRIE members

GISAID acknowledgements

## Data Availability

There are no new sequence data in this work

## Acknowledgements

We thank Russell Corbett-Detig and Yatish Turakhia for invaluable help with UShER, and Alexey Kondrashov for valuable discussions. We are grateful to all GISAID submitting and originating labs (Supplementary File 3) for rapid open release of SARS-CoV-2 sequencing data.

NS and VS were funded within the framework of the HSE University Basic Research Program.

## References

[1] World Health Organization. Tracking SARS-CoV-2 variants n.d.

[2] Davies NG, Abbott S, Barnard RC, Jarvis CI, Kucharski AJ, Munday JD, et al. Estimated transmissibility and impact of SARS-CoV-2 lineage B.1.1.7 in England. Science 2021;372:eabg3055. https://doi.org/10.1126/science.abg3055.

[3] Endo A, Centre for the Mathematical Modelling of Infectious Diseases COVID-19 Working Group, Abbott S, Kucharski AJ, Funk S. Estimating the overdispersion in COVID-19 transmission using outbreak sizes outside China. Wellcome Open Res 2020;5:67. https://doi.org/10.12688/wellcomeopenres.15842.3.

[4] Lewis D. Superspreading drives the COVID pandemic - and could help to tame it. Nature 2021;590:544–6. https://doi.org/10.1038/d41586-021-00460-x.

[5] Sun K, Wang W, Gao L, Wang Y, Luo K, Ren L, et al. Transmission heterogeneities, kinetics, and controllability of SARS-CoV-2. Science 2021;371:eabe2424. https://doi.org/10.1126/science.abe2424.

[6] Mlcochova P, Kemp SA, Dhar MS, Papa G, Meng B, Ferreira IATM, et al. SARS-CoV-2 B.1.617.2 Delta variant replication and immune evasion. Nature 2021;599:114–9. https://doi.org/10.1038/s41586-021-03944-y.

[7] Hodcroft EB. CoVariants: SARS-CoV-2 Mutations and Variants of Interest 2021.

[8] Fisman DN, Tuite AR. Evaluation of the relative virulence of novel SARS-CoV-2 variants: a retrospective cohort study in Ontario, Canada. CMAJ 2021;193:E1619–25. https://doi.org/10.1503/cmaj.211248.

[9] Li M, Lou F, Fan H. SARS-CoV-2 Variants of Concern Delta: a great challenge to prevention and control of COVID-19. Signal Transduct Target Ther 2021;6:349. https://doi.org/10.1038/s41392-021-00767-1.

[10] Planas D, Veyer D, Baidaliuk A, Staropoli I, Guivel-Benhassine F, Rajah MM, et al. Reduced sensitivity of SARS-CoV-2 variant Delta to antibody neutralization. Nature 2021;596:276–80. https://doi.org/10.1038/s41586-021-03777-9.

[11] Arora P, Sidarovich A, Krüger N, Kempf A, Nehlmeier I, Graichen L, et al. B.1.617.2 enters and fuses lung cells with increased efficiency and evades antibodies induced by infection and vaccination. Cell Rep 2021;37:109825. https://doi.org/10.1016/j.celrep.2021.109825.

[12] Borisova NI, Kotov IA, Kolesnikov AA, Kaptelova VV, Speranskaya AS, Kondrasheva LY, et al. [Monitoring the spread of the SARS-CoV-2 (Coronaviridae: Coronavirinae: Betacoronavirus; Sarbecovirus) variants in the Moscow region using targeted high-throughput sequencing]. Vopr Virusol 2021;66:269–78. https://doi.org/10.36233/0507-4088-72.

[13] Dmitry D. Knorre, Elena Nabieva, Sofya K. Garushyants, The CoRGI (Coronavirus Russian Genetic Initiative) Consortium, and Georgii A. Bazykin. taxameter.ru. Available online: http://taxameter.ru/ (2021) n.d.

[14] Stern A, Fleishon S, Kustin T, Dotan E, Mandelboim M, Erster O, et al. The unique evolutionary dynamics of the SARS-CoV-2 Delta variant. Infectious Diseases (except HIV/AIDS); 2021. https://doi.org/10.1101/2021.08.05.21261642.

[15] Chadeau-Hyam M, Eales O, Bodinier B, Wang H, Haw D, Whitaker M, et al. REACT-1 round 15 interim report: High and rising prevalence of SARS-CoV-2 infection in England from end of September 2021 followed by a fall in late October 2021. Epidemiology; 2021. https://doi.org/10.1101/2021.11.03.21265877.

[16] Arora P, Kempf A, Nehlmeier I, Graichen L, Sidarovich A, Winkler MS, et al. Delta variant (B.1.617.2) sublineages do not show increased neutralization resistance. Cell Mol Immunol 2021;18:2557–9. https://doi.org/10.1038/s41423-021-00772-y.

[17] UK Health Security Agency. Technical briefing 28; SARS-CoV-2 variants of concern and variants under investigation in England 2021.

[18] Kistler KE, Huddleston J, Bedford T. Rapid and parallel adaptive mutations in spike S1 drive clade success in SARS-CoV-2. BioRxiv 2021:2021.09.11.459844. https://doi.org/10.1101/2021.09.11.459844.

[19] Turakhia Y, Thornlow B, Hinrichs AS, De Maio N, Gozashti L, Lanfear R, et al. Ultrafast Sample placement on Existing tRees (UShER) enables real-time phylogenetics for the SARS-CoV-2 pandemic. Nat Genet 2021;53:809–16. https://doi.org/10.1038/s41588-021-00862-7.

[20] McBroome J, Thornlow B, Hinrichs AS, Kramer A, De Maio N, Goldman N, et al. A Daily-Updated Database and Tools for Comprehensive SARS-CoV-2 Mutation-Annotated Trees. Molecular Biology and Evolution 2021:msab264. https://doi.org/10.1093/molbev/msab264.

[21] Price MN, Dehal PS, Arkin AP. FastTree 2 – Approximately Maximum-Likelihood Trees for Large Alignments. PLoS ONE 2010;5:e9490. https://doi.org/10.1371/journal.pone.0009490.

[22] R Core Team. R: A language and environment for statistical computing 2021.

[23] Baty F, Ritz C, Charles S, Brutsche M, Flandrois J-P, Delignette-Muller M-L. A Toolbox for Nonlinear Regression in R: The Package nlstools. Journal of Statistical Software 2015:1–21.

[24] Stadler T, Kühnert D, Bonhoeffer S, Drummond AJ. Birth-death skyline plot reveals temporal changes of epidemic spread in HIV and hepatitis C virus (HCV). Proc Natl Acad Sci U S A 2013;110:228–33. https://doi.org/10.1073/pnas.1207965110.

[25] Bouckaert R, Vaughan TG, Barido-Sottani J, Duchêne S, Fourment M, Gavryushkina A, et al. BEAST 2.5: An advanced software platform for Bayesian evolutionary analysis. PLoS Comput Biol 2019;15:e1006650. https://doi.org/10.1371/journal.pcbi.1006650.

[26] Rambaut A, Drummond AJ, Xie D, Baele G, Suchard MA. Posterior Summarization in Bayesian Phylogenetics Using Tracer 1.7. Systematic Biology 2018;67:901–4. https://doi.org/10.1093/sysbio/syy032.

[27] Wickham H, Averick M, Bryan J, Chang W, McGowan L, François R, et al. Welcome to the Tidyverse. JOSS 2019;4:1686. https://doi.org/10.21105/joss.01686.

[28] Slowikowski K. ggrepel: Automatically Position Non-Overlapping Text Labels with “ggplot2”. R package version 0.9.1. 2021.

[29] Auguie B. egg: Extensions for “ggplot2”: Custom Geom, Custom Themes, Plot Alignment, Labelled Panels, Symmetric Scales, and Fixed Panel Size. R package version 0.4.5 2019.

[30] Wickham H. stringr: Simple, Consistent Wrappers for Common String Operations. R package version 1.4.0 2019.

[31] Harrell Jr FE. Hmisc: Harrell Miscellaneous. R package version 4.6-0 2021.

[32] Huerta-Cepas J, Serra F, Bork P. ETE 3: Reconstruction, Analysis, and Visualization of Phylogenomic Data. Mol Biol Evol 2016;33:1635–8. https://doi.org/10.1093/molbev/msw046.

[33] Public Health England. SARS-CoV-2 variants of concern and variants under investigation in England; Technical briefing 15 2021.

[34] Chen C, Nadeau SA, Yared M, Voinov P, Stadler T. CoV-Spectrum: Analysis of globally shared SARS-CoV-2 data to Identify and Characterize New Variants. ArXiv:210608106 [q-Bio] 2021.

[35] Sagulenko P, Puller V, Neher RA. TreeTime: Maximum-likelihood phylodynamic analysis. Virus Evolution 2018;4. https://doi.org/10.1093/ve/vex042.

[36] Matsvay A, Klink GV, Safina KR, Nabieva E, Garushyants SK, Biba D, et al. Genomic epidemiology of SARS-CoV-2 in Russia reveals recurring cross-border transmission throughout 2020. Epidemiology; 2021. https://doi.org/10.1101/2021.03.31.21254115.

[37] Gupta M, Azumaya CM, Moritz M, Pourmal S, Diallo A, Merz GE, et al. CryoEM and AI reveal a structure of SARS-CoV-2 Nsp2, a multifunctional protein involved in key host processes. Biophysics; 2021. https://doi.org/10.1101/2021.05.10.443524.

[38] Kosakovsky Pond SL, Poon AFY, Velazquez R, Weaver S, Hepler NL, Murrell B, et al. HyPhy 2.5—A Customizable Platform for Evolutionary Hypothesis Testing Using Phylogenies. Molecular Biology and Evolution 2020;37:295–9. https://doi.org/10.1093/molbev/msz197.

[39] Martin-Sancho L, Lewinski MK, Pache L, Stoneham CA, Yin X, Becker ME, et al. Functional landscape of SARS-CoV-2 cellular restriction. Molecular Cell 2021;81:2656–2668.e8. https://doi.org/10.1016/j.molcel.2021.04.008.

[40] Zhou Z, Huang C, Zhou Z, Huang Z, Su L, Kang S, et al. Structural insight reveals SARS-CoV-2 ORF7a as an immunomodulating factor for human CD14+ monocytes. IScience 2021;24:102187. https://doi.org/10.1016/j.isci.2021.102187.

[41] Nemudryi A, Nemudraia A, Wiegand T, Nichols J, Snyder DT, Hedges JF, et al. SARS-CoV-2 genomic surveillance identifies naturally occurring truncation of ORF7a that limits immune suppression. Cell Reports 2021;35:109197. https://doi.org/10.1016/j.celrep.2021.109197.

[42] Foster CSP, Rawlinson WD. Rapid spread of a SARS-CoV-2 Delta variant with a frameshift deletion in ORF7a. Genetic and Genomic Medicine; 2021. https://doi.org/10.1101/2021.08.18.21262089.

[43] Moody R, Wilson KL, Boer JC, Holien JK, Flanagan KL, Jaworowski A, et al. Predicted B Cell Epitopes Highlight the Potential for COVID-19 to Drive Self-Reactive Immunity. Front Bioinform 2021;1:709533. https://doi.org/10.3389/fbinf.2021.709533.

[44] Kozlovskaya L, Piniaeva A, Ignatyev G, Selivanov A, Shishova A, Kovpak A, et al. Isolation and phylogenetic analysis of SARS-CoV-2 variants collected in Russia during the COVID-19 outbreak. International Journal of Infectious Diseases 2020;99:40–6. https://doi.org/10.1016/j.ijid.2020.07.024.

[45] Komissarov AB, Safina KR, Garushyants SK, Fadeev AV, Sergeeva MV, Ivanova AA, et al. Genomic epidemiology of the early stages of the SARS-CoV-2 outbreak in Russia. Nat Commun 2021;12:649. https://doi.org/10.1038/s41467-020-20880-z.

[46] Klink GV, Safina KR, Garushyants SK, Moldovan M, Nabieva E, The CoRGI (Coronavirus Russian Genetic Initiative) Consortium, et al. Spread of endemic SARS-CoV-2 lineages in Russia. Epidemiology; 2021. https://doi.org/10.1101/2021.05.25.21257695.

[47] Hodcroft EB, Zuber M, Nadeau S, Vaughan TG, Crawford KHD, Althaus CL, et al. Spread of a SARS-CoV-2 variant through Europe in the summer of 2020. Nature 2021;595:707–12. https://doi.org/10.1038/s41586-021-03677-y.

[48] Ruan Y, Luo Z, Tang X, Li G, Wen H, He X, et al. On the founder effect in COVID-19 outbreaks: how many infected travelers may have started them all? National Science Review 2021;8:nwaa246. https://doi.org/10.1093/nsr/nwaa246.

