## Supplementary tables and figures for "The rise and spread of the SARS-CoV-2 AY.122 lineage in Russia"

### Supplementary Materials

**Table S1. The priors used for the BDSKY model for phylodynamic analysis.** Clock rate was set according to the previously estimated value in [1]. Becoming uninfectious rate corresponds to an average of 7 days being infectious [2]

| Parameter | Prior |
|---|---|
| clockModel | Strict clock with fixed mean (0.0009) |
| Re change times | '2021-09-01 2021-08-01 2021-07-01 2021-06-01 2021-05-01' |
| sampling proportion change times | '2021-09-01 2021-08-01 2021-07-01 2021-06-01 2021-05-01' |
| becoming uninfectious rate | 52.2857 (fixed) |

**Table S2.** Fractions of samples belonging to the main import among Delta samples from different regions of Russia. Regions with more than five samples of Delta lineage are shown in bold.

| region | Delta samples | earliest Delta sample | main import samples | earliest main import sample | fraction of main import |
|---|---|---|---|---|---|
| Bashkortostan | 4 | 2021-06-15 | 3 | 2021-06-15 | 0.75 |
| **Belgorod** | **13** | **2021-07-16** | **13** | **2021-07-16** | **1** |
| Bryansk | 1 | 2021-07-05 | 1 | 2021-07-05 | 1 |
| **Buryatia** | **6** | **2021-07-01** | **6** | **2021-07-01** | **1** |
| Chelyabinsk | 3 | 2021-06-27 | 3 | 2021-06-27 | 1 |
| **Chukotka** | **8** | **2021-06-17** | **8** | **2021-06-17** | **1** |
| **Ivanovo** | **24** | **2021-06-11** | **21** | **2021-06-11** | **0.88** |
| Kabardino-Balkaria | 1 | 2021-07-05 | 1 | 2021-07-05 | 1 |
| Kaliningrad | 1 | 2021-06-09 | 1 | 2021-06-09 | 1 |
| **Kamchatka** | **17** | **2021-06-15** | **16** | **2021-06-15** | **0.94** |
| **Karachay-Cherkess** | **25** | **2021-06-09** | **24** | **2021-06-09** | **0.96** |
| **Kemerovo** | **6** | **2021-06-16** | **5** | **2021-06-16** | **0.83** |
| **Khanty-Mansi** | **6** | **2021-06-16** | **4** | **2021-06-16** | **0.67** |
| Kirov | 2 | 2021-06-15 | 2 | 2021-06-15 | 1 |
| **Kostroma** | **30** | **2021-06-14** | **29** | **2021-06-14** | **0.97** |
| **Krasnodar** | **31** | **2021-06-09** | **30** | **2021-06-09** | **0.97** |
| Kursk | 2 | 2021-06-15 | 2 | 2021-06-15 | 1 |
| Lipetsk | 3 | 2021-07-02 | 3 | 2021-07-02 | 1 |
| **Magadan** | **19** | **2021-06-14** | **18** | **2021-06-14** | **0.95** |

| | | | | | |
|---|---|---|---|---|---|
| **Moscow** | **637** | **2021-04-19** | **573** | **2021-04-19** | **0.9** |
| Murmansk | 2 | 2021-07-05 | 2 | 2021-07-05 | 1 |
| Novgorod | 4 | 2021-06-22 | 4 | 2021-06-22 | 1 |
| Orenburg | 1 | 2021-07-05 | 1 | 2021-07-05 | 1 |
| Penza | 5 | 2021-05-23 | 3 | 2021-05-23 | 0.6 |
| **Perm** | **6** | **2021-07-10** | **5** | **2021-07-10** | **0.83** |
| Pskov | 1 | 2021-07-04 | 1 | 2021-07-04 | 1 |
| **Rostov** | **6** | **2021-07-05** | **5** | **2021-07-05** | **0.83** |
| Ryazan | 3 | 2021-06-29 | 2 | 2021-06-29 | 0.67 |
| **Saint-Petersburg** | **330** | **2021-04-12** | **304** | **2021-04-23** | **0.92** |
| **Saratov** | **7** | **2021-06-11** | **7** | **2021-06-11** | **1** |
| **Simferopol** | **8** | **2021-07-01** | **6** | **2021-07-01** | **0.75** |
| **Smolensk** | **20** | **2021-07-06** | **18** | **2021-07-07** | **0.9** |
| **Tambov** | **17** | **2021-06-10** | **17** | **2021-06-10** | **1** |
| **Tver** | **16** | **2021-04-07** | **10** | **2021-06-15** | **0.625** |
| **Tyumen** | **7** | **2021-06-22** | **4** | **2021-06-22** | **0.57** |
| **Ulyanovsk** | **7** | **2021-06-15** | **7** | **2021-06-15** | **1** |
| **Vladimir** | **90** | **2021-06-15** | **89** | **2021-06-15** | **0.99** |
| **Vologda** | **8** | **2021-06-14** | **6** | **2021-06-15** | **0.75** |
| Voronezh | 3 | 2021-06-09 | 3 | 2021-06-09 | 1 |
| **Yaroslavl** | **48** | **2021-05-06** | **46** | **2021-05-06** | **0.96** |
| **Zabaykalsky** | **6** | **2021-06-09** | **5** | **2021-06-09** | **0.83** |

**Table S3.** Number of GISAID samples retained after filtering (see Methods) for countries shown in Figure 5.

| **Country** | **Filtered samples** |
|---|---|
| Australia | 8722 |
| Austria | 2526 |
| Belgium | 13509 |
| Brazil | 8273 |
| Canada | 41244 |
| Croatia | 2123 |
| Czech Republic | 2721 |
| Denmark | 50910 |
| Finland | 5835 |
| France | 34531 |
| Germany | 59222 |
| Greece | 1834 |
| Iceland | 3383 |
| India | 17059 |
| Indonesia | 2924 |
| Ireland | 12950 |
| Italy | 18703 |
| Japan | 40597 |
| Lithuania | 3747 |

| | |
|---|---|
| Malaysia | 1795 |
| Mexico | 9150 |
| Netherlands | 18782 |
| Norway | 8089 |
| Poland | 2764 |
| Portugal | 7394 |
| Romania | 2108 |
| Russia | 1440 |
| Singapore | 5840 |
| Slovakia | 2789 |
| Slovenia | 7953 |
| South Africa | 3802 |
| South Korea | 4306 |
| Spain | 14114 |
| Sweden | 22967 |
| Switzerland | 20058 |
| Turkey | 44690 |
| United Kingdom | 472589 |
| USA | 443374 |

**Table S4.** Dates of the first sample with the nsp2:K81N+ORF7a:P45L combination of mutations in each country where it has been observed. Countries where this variant was detected before Russia are shown in bold. Countries that are among ten locations with the highest passenger traffic with Russia in the first half of 2021 are underlined.

| Country | Date of earliest sample |
|---|---|
| **Sweden*** | **2020-11-19** |
| **Slovakia**** | **2021-01-09** |
| **Italy***** | **2021-02-07** |
| **Slovenia****** | **2021-02-10** |
| **Czech Republic******* | **2021-02-12** |
| **Turkey********** | **2021-03-09** |
| **Israel** | **2021-04-10** |
| **Singapore** | **2021-04-15** |
| **Japan** | **2021-04-16** |
| **Switzerland** | **2021-04-17** |

| | |
|---|---|
| USA | 2021-04-19 |
| Northern Ireland | 2021-04-22 |
| England | 2021-04-22 |
| Germany | 2021-04-26 |
| Austria | 2021-04-26 |
| Georgia | 2021-05-12 |
| Poland | 2021-05-14 |
| Latvia | 2021-05-15 |
| France | 2021-05-17 |
| Netherlands | 2021-05-20 |
| Monaco | 2021-05-21 |
| Lithuania | 2021-05-27 |
| Norway | 2021-05-27 |

| | |
|---|---|
| Spain | 2021-05-28 |
| Denmark | 2021-05-31 |
| Iceland | 2021-06-01 |
| Scotland | 2021-06-02 |
| Luxembourg | 2021-06-02 |
| Ireland | 2021-06-05 |
| New Zealand | 2021-06-06 |
| Canada | 2021-06-07 |
| Portugal | 2021-06-08 |
| Finland | 2021-06-09 |
| Belgium | 2021-06-10 |
| Estonia | 2021-06-10 |
| Wales | 2021-06-11 |

| | |
|---|---|
| South Korea | 2021-06-12 |
| Chile | 2021-06-17 |
| Greece | 2021-06-17 |
| Moldova | 2021-06-17 |
| Ukraine | 2021-06-18 |
| Montenegro | 2021-06-18 |
| Hong Kong | 2021-06-19 |
| Brazil | 2021-06-21 |
| Bulgaria | 2021-06-21 |
| Ecuador | 2021-06-22 |
| Malawi | 2021-06-22 |
| Croatia | 2021-06-25 |
| Uzbekistan | 2021-06-25 |

| | |
|---|---|
| Serbia | 2021-06-26 |
| Mexico | 2021-06-28 |
| Curacao | 2021-06-28 |
| Bosnia and Herzegovina | 2021-06-30 |
| Romania | 2021-07-02 |
| Bahrain | 2021-07-02 |
| Guatemala | 2021-07-05 |
| Australia | 2021-07-08 |
| Peru | 2021-07-10 |
| Sint Maarten | 2021-07-19 |
| Guadeloupe | 2021-07-20 |
| North Macedonia | 2021-07-20 |
| Jiangsu | 2021-07-20 |

| | |
|---|---|
| Costa Rica | 2021-07-21 |
| Puerto Rico | 2021-07-21 |
| Kosovo | 2021-07-25 |
| Colombia | 2021-07-26 |
| Gibraltar | 2021-07-28 |
| Argentina | 2021-08-15 |
| Thailand | 2021-08-16 |
| India | 2021-08-17 |
| Kazakhstan | 2021-08-17 |
| Nigeria | 2021-08-26 |
| Vietnam | 2021-09-03 |
| Ghana | 2021-09-04 |
| Indonesia | 2021-09-05 |

| | |
|---|---|
| Liechtenstein | 2021-09-07 |
| Aruba | 2021-09-13 |

* submitted on August 30, 2021; the next sample with this combination of mutations in Sweden dates to May, 2021.

** probably erroneous dates (likely confused month and day: samples carrying this mutation date to Jan 9, Feb 9, March 9 and April 9, and the opposite ordering of month and day to that in GISAID submission forms is common in Europe).

*** submitted on July 20, 2021; the next sample with this pair of mutations in Italy dates to May 20, 2021.

**** submitted on October 19, 2021; the next sample with this pair of mutations in Slovenia dates to June 5, 2021.

***** submitted to GISAID on August 10, 2021; the next sample with this pair of mutations in Czech Republic dates to April 22, 2021.

******submitted to GISAID on September 09, 2021; the next sample with this pair of mutations in Turkey dates to May 24, 2021.

**Table S5.** Traffic volumes through Russian airports for international airlines in 2020-2021 according to Federal Air Transport Agency of Russia (https://favt.gov.ru/dejatelnost-ajeroporty-i-ajerodromy-osnovnie-proizvodstvennie-pokazateli-aeroportov-obyom-perevoz/)

| **year** | **month** | **Passenger traffic: departing + arriving passengers** |
|---|---|---|
| 2020 | 1 | 4 929 170 |
| 2020 | 2 | 4 166 359 |
| 2020 | 3 | 2 505 968 |
| 2020 | 4 | 30 426 |
| 2020 | 5 | 48 884 |
| 2020 | 6 | 69 601 |
| 2020 | 7 | 110 828 |
| 2020 | 8 | 912 802 |
| 2020 | 9 | 1 522 681 |
| 2020 | 10 | 1 430 033 |
| 2020 | 11 | 720 836 |
| 2020 | 12 | 731 440 |
| 2021 | 1 | 727 361 |
| 2021 | 2 | 756 012 |
| 2021 | 3 | 1 205 488 |
| 2021 | 4 | 1 346 025 |
| 2021 | 5 | 1 452 228 |
| 2021 | 6 | 2 095 960 |

| | | |
|---|---|---|
| 2021 | 7 | 3 464 726 |
| 2021 | 8 | 4 023 019 |
| 2021 | 9 | 4 138 497 |

**Figure S1. Phylogenetic inference of imports into Russia.** Tips are marked as Russian (*R*) or non-Russian (*O*) by place of collection. All internal nodes are numbered in order along each lineage from root to tip. Moving from the nodes with the highest numbers (here, 4) towards the lowest (root), each node N is labelled according to the labels of its immediate descendants (tips or internal nodes) as follows. If more than one descendant is labelled *R*, N is labelled *R*. If no descendants are labelled *R*, N is not labelled. If exactly one descendant is labelled *R*, the branch leading to this descendant is marked as an import, and N is not labelled. In case of nested imports, only the deepest import marks are retained; i.e., among multiple imports inferred for a root-to-tip lineage, only the one closest to the root is retained. In the illustrated case, only the dark purple import is retained. Imports into other countries were identified analogously.

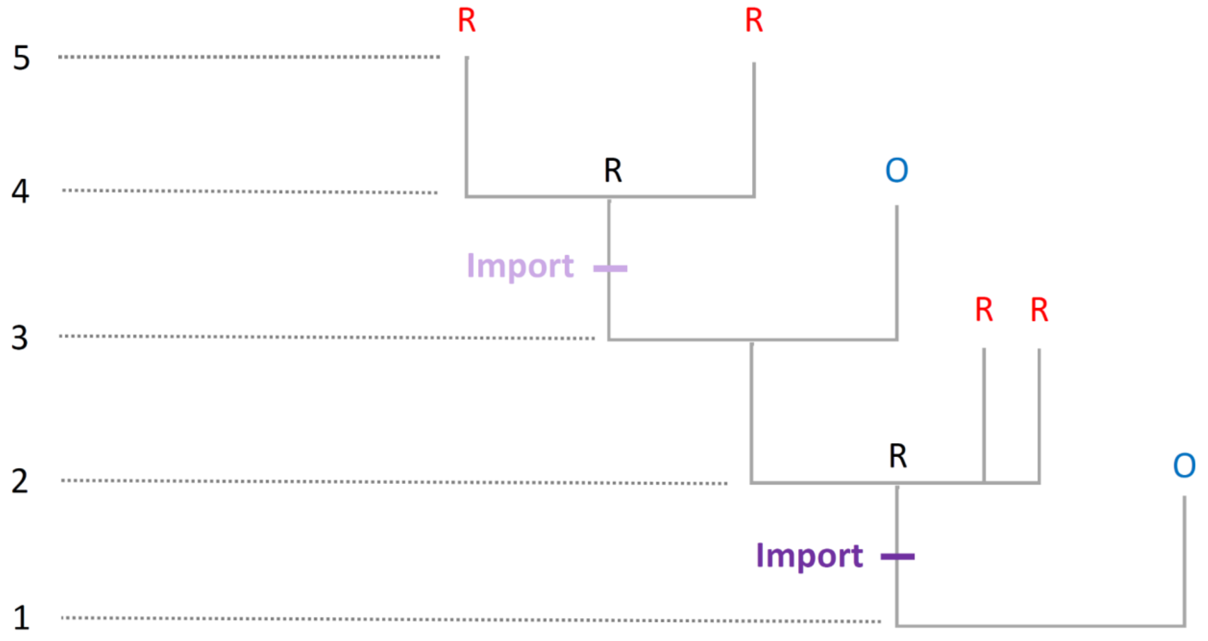

**Figure S2. The effect of subsampling on the number of samples used for the phylodynamic analysis**

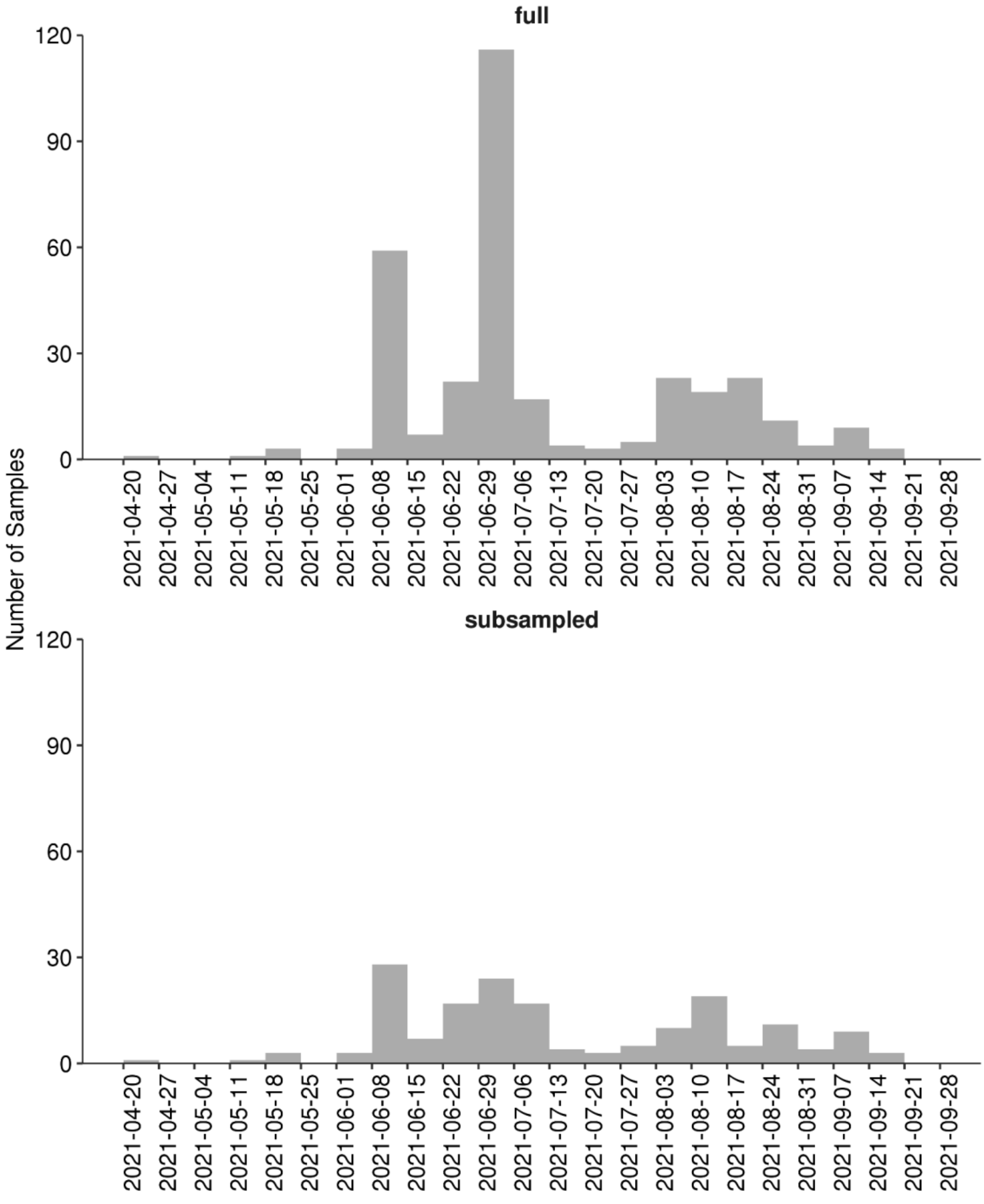

**Figure S3**. **UpSet plot for mutations of the Delta lineage with frequency greater than 5% in Russia.** Only the combinations that were seen in more than five samples in Russia are shown. The following mutations that characterize the major sublineage of B.1.617.2 ("21J" in Nextstrain nomenclature) and occur in >85% of Delta samples both in Russia and globally are not shown: RdRp:G671S, exonuclease:A394V, nsp6:T77A, nsp3:A488S, nsp3:P1228L, nsp6:V120V, ORF7b:T40I, nsp3:P1469S, N:G215C, nsp4:D144D, nsp4:V167L, and nsp4:T492I.

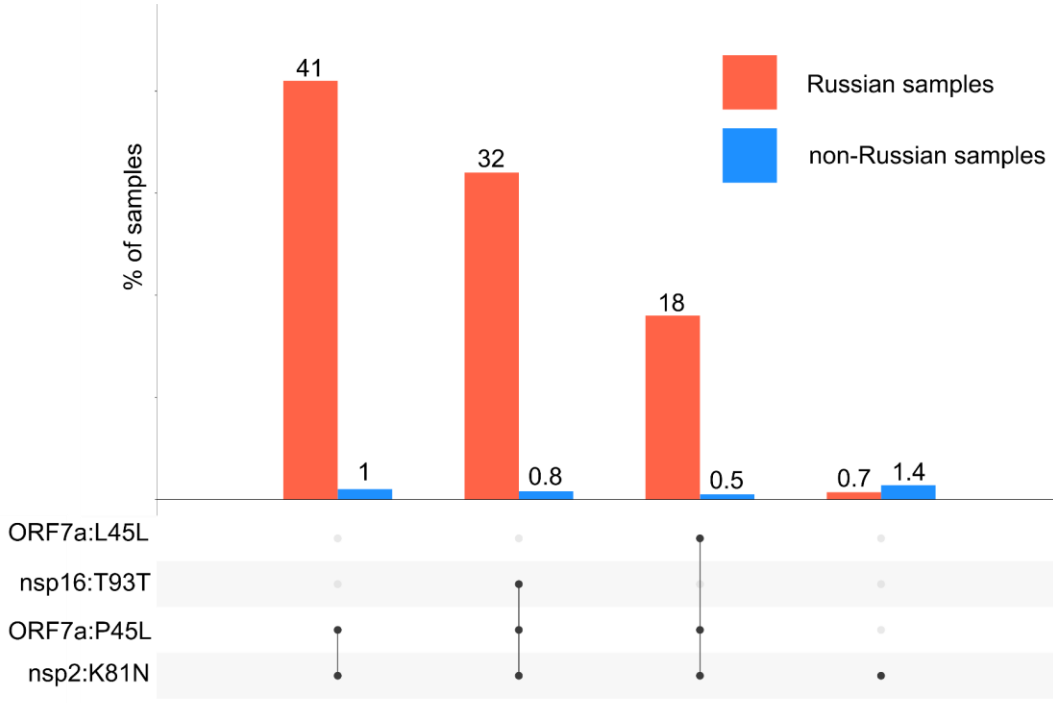

**Figure S4. Timeline for imports of Delta subclades into Russia's regions.** The figure is similar to Fig. 3B, but the samples from Moscow (A), Saint Petersburg (B) and other regions of Russia (C) are shown separately.

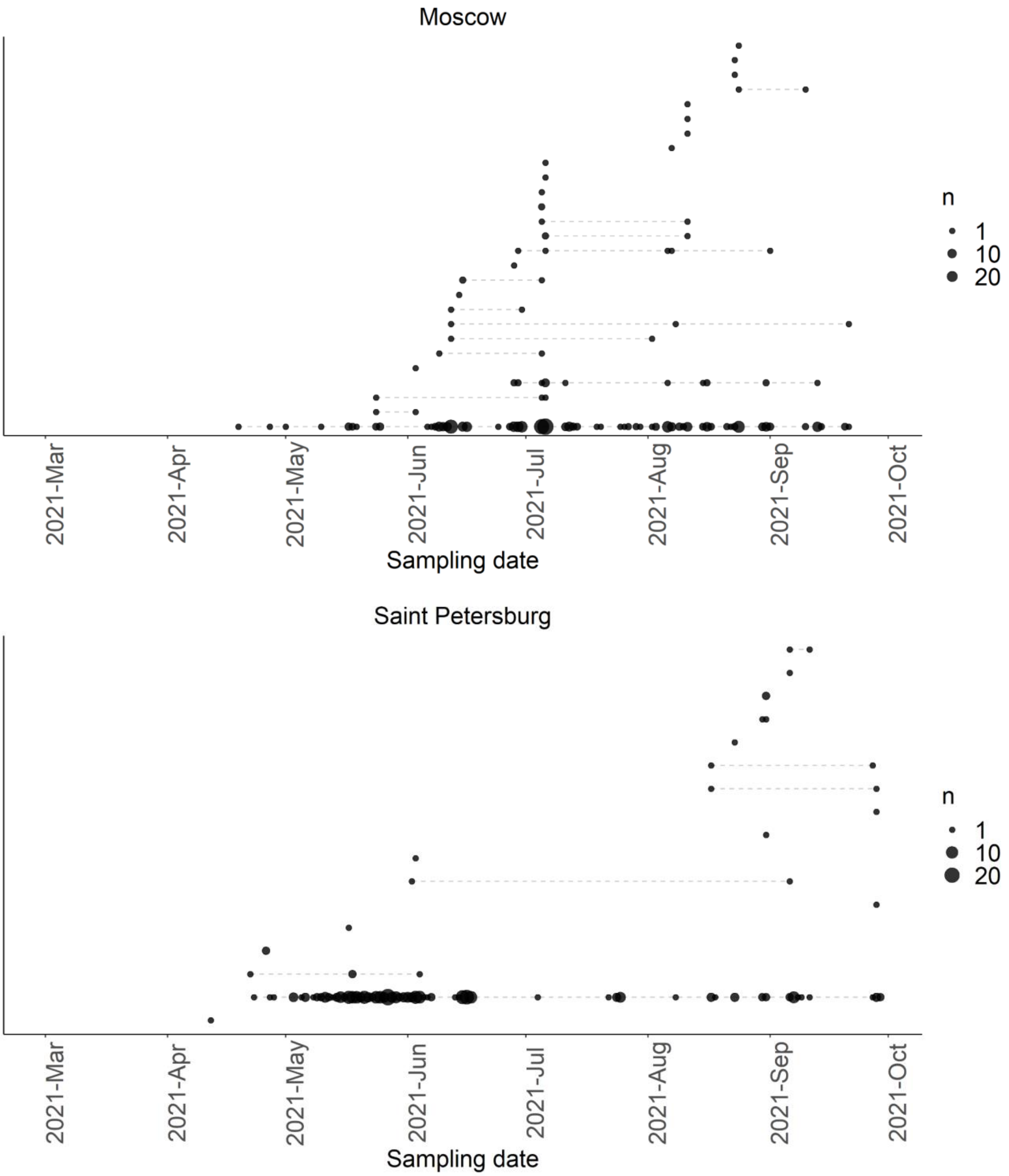

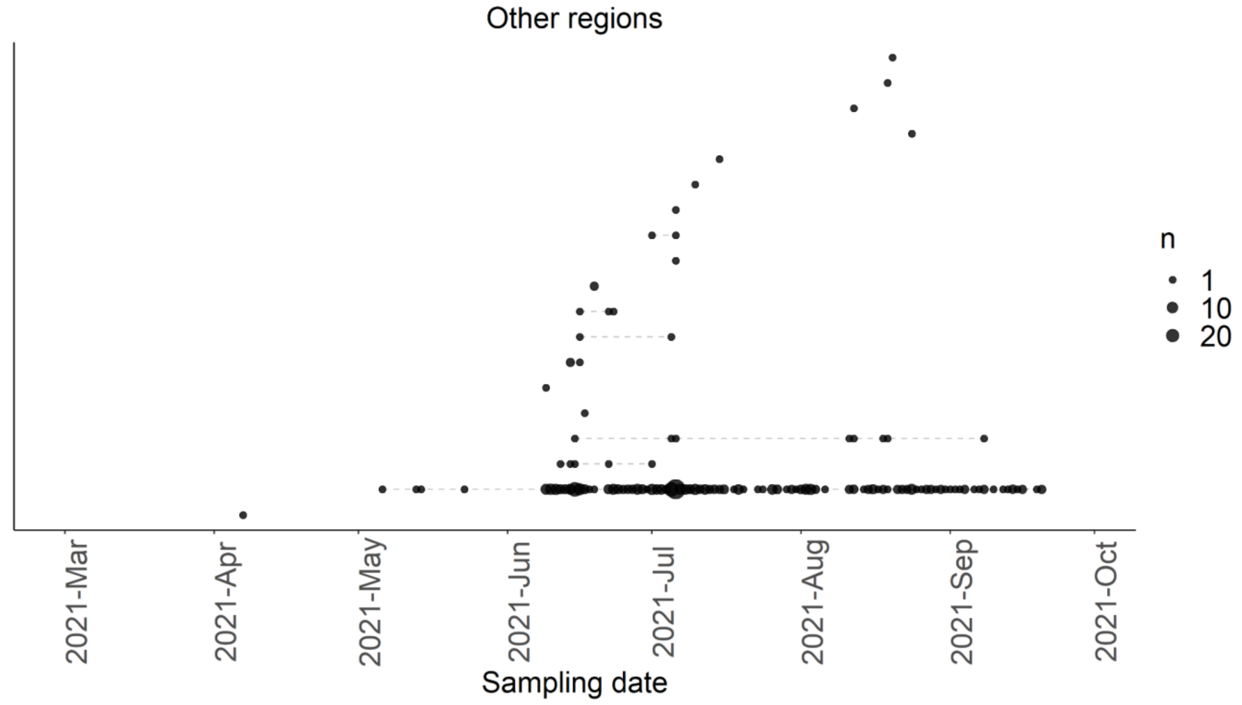
Other regions
n
1
10
20
2021-Mar
2021-Apr
2021-May
2021-Jun
2021-Jul
2021-Aug
2021-Sep
2021-Oct
Sampling date

**Figure S5. Frequency of the ORF7a:P45L variant among all Russian samples by month of sampling.**

April 2021 (https://taxameter.ru/?mutation=ORF7a:P45L&lang=EN&min_date=2021-4-1&max_date=2021-4-30)

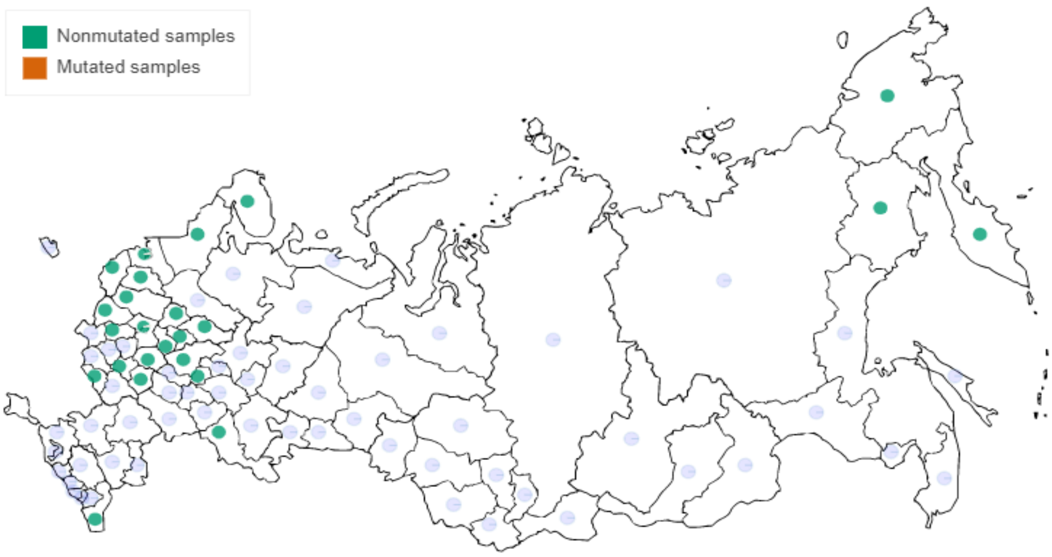


May 2021 (https://taxameter.ru/?mutation=ORF7a:P45L&lang=EN&min_date=2021-5-1&max_date=2021-5-31)

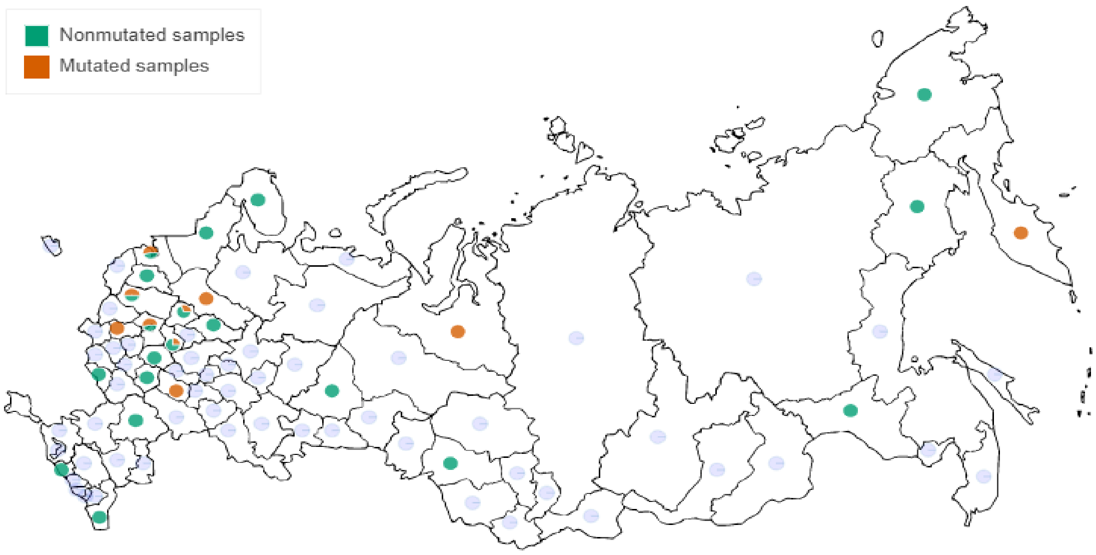

June 2021 (https://taxameter.ru/?mutation=ORF7a:P45L&lang=EN&min_date=2021-6-1&max_date=2021-6-30)

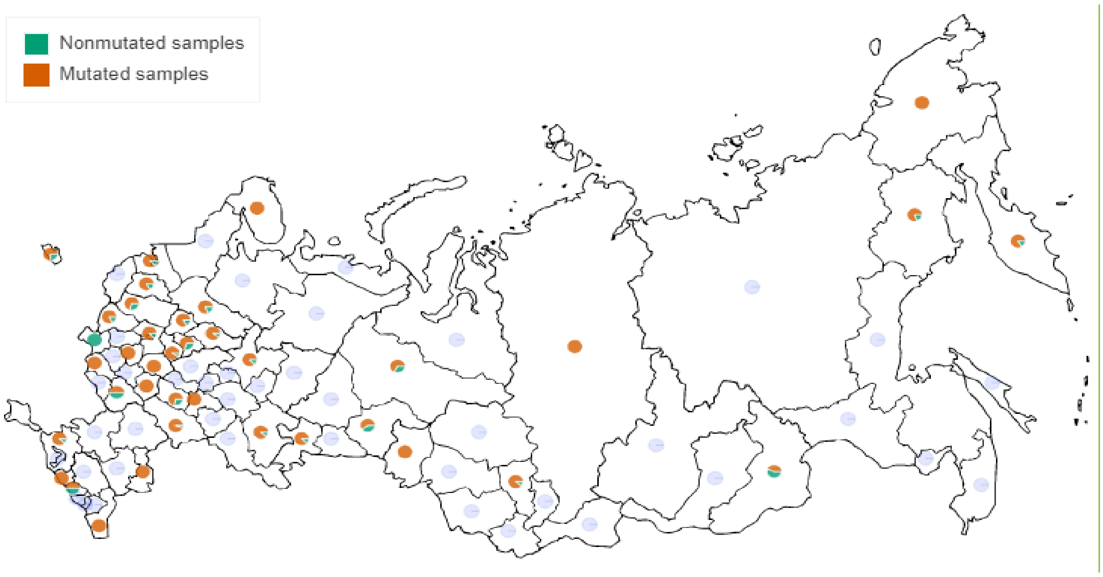


July-October (up to Oct 13, https://taxameter.ru/?mutation=ORF7a:P45L&lang=EN&min_date=2021-7-1&max_date=2021-10-13)

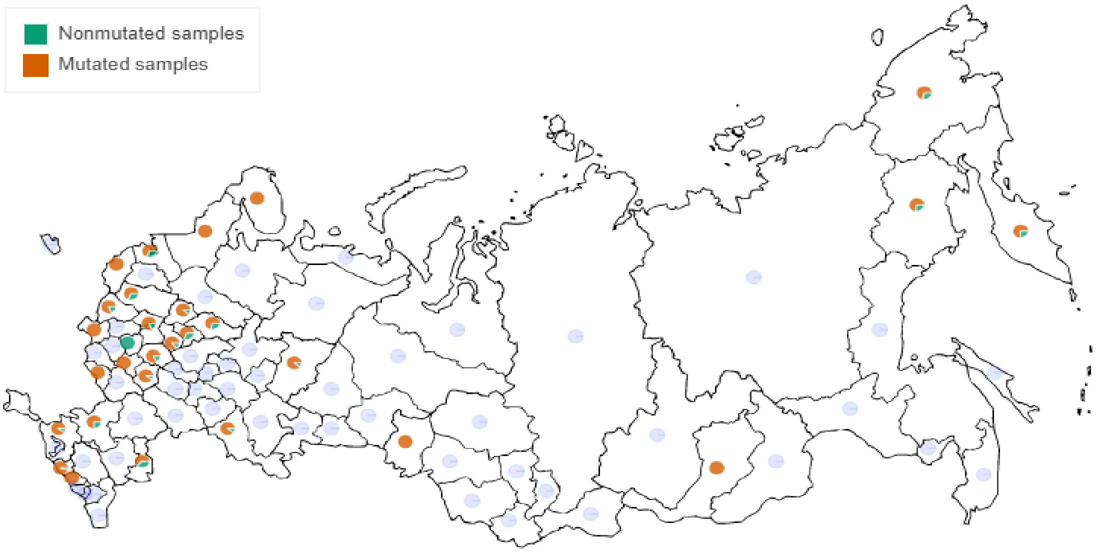

**Figure S6. Re inference is robust to the density of subsampling of overrepresented dates.**

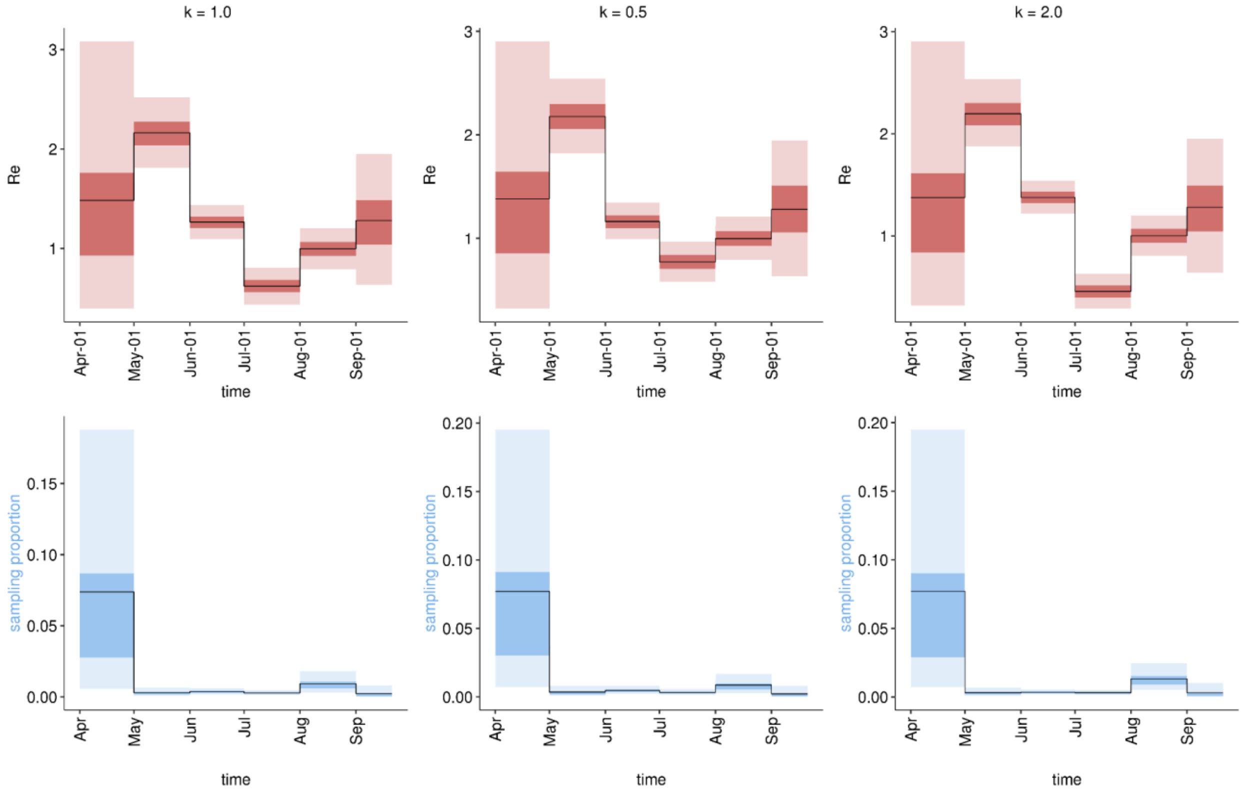

**Figure S7. Dynamics of nsp2:K81N+ORF7a:P45L frequency among all samples in various countries**. A) Frequencies of nsp2:K81N+ORF7a:P45L among ALL samples measured in 15-days sliding windows; windows with less than 20 samples are filtered out. B) Logistic growth rates estimated for nsp2:K81N+ORF7a:P45L among Delta samples before and after July 1st.

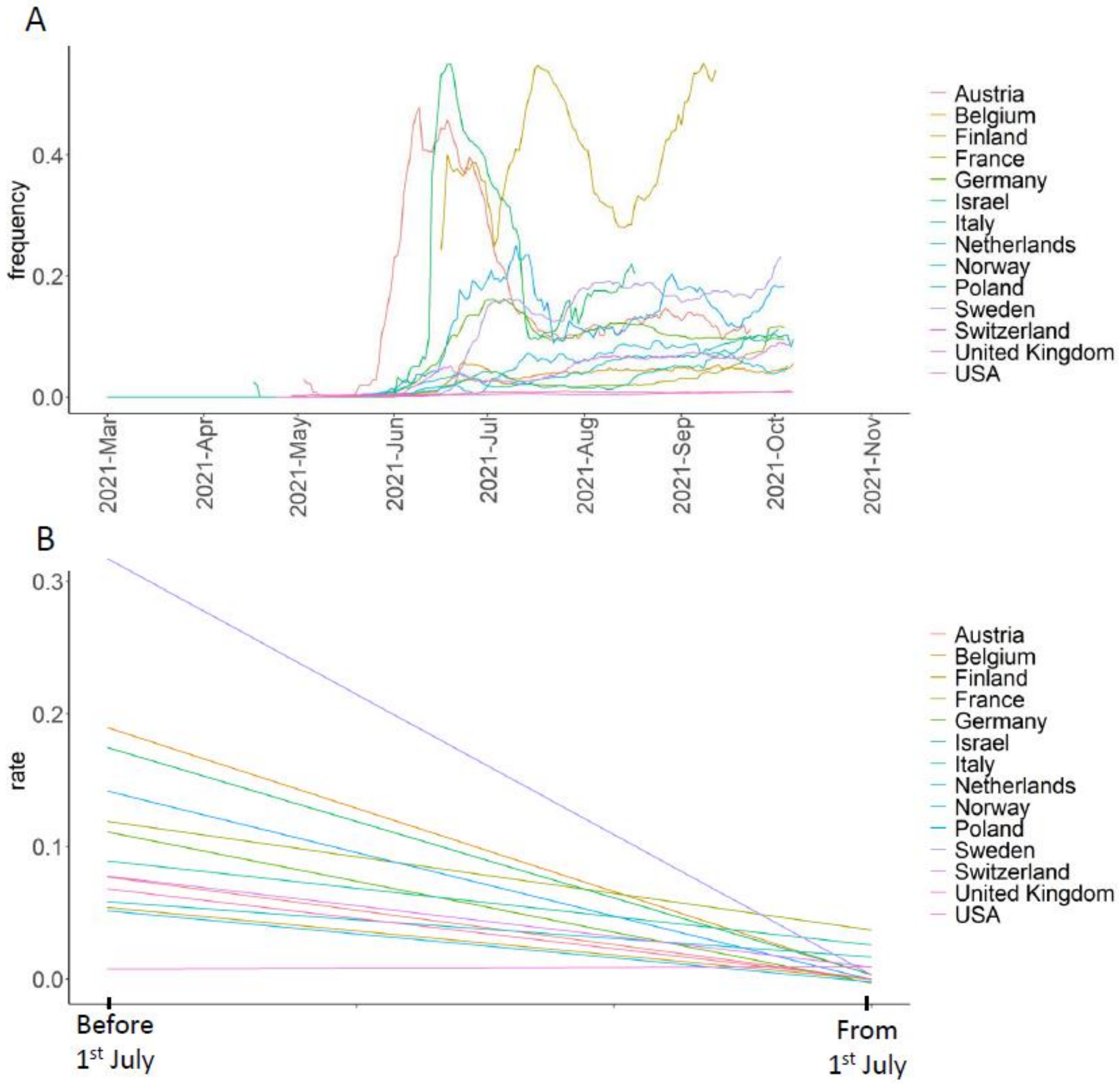

**Figure S8. Dynamics of nsp2:K81N+ORF7a:P45L frequency among Delta samples in various countries**. A) Frequencies of nsp2:K81N+ORF7a:P45L among Delta samples measured in 15-days sliding windows; windows with less than 20 Delta samples are filtered out. B) Logistic growth rates estimated for nsp2:K81N+ORF7a:P45L among Delta samples before and from July 1st.

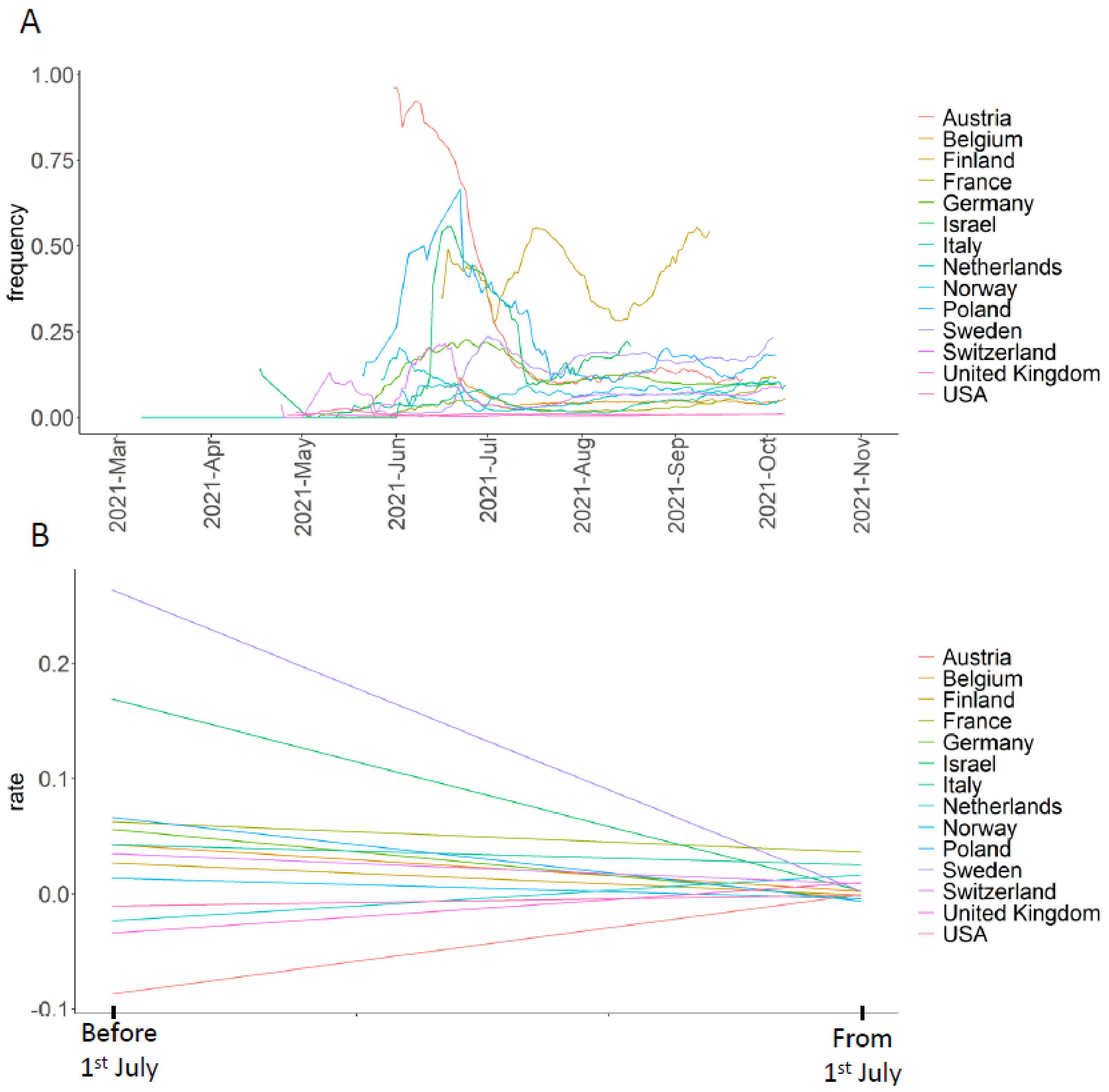

**Figure S9. Fraction of Delta samples in the largest import and relatedness of Delta samples, for countries with at least 50 Delta samples in the UShER tree.** Notation as in Fig. 5.

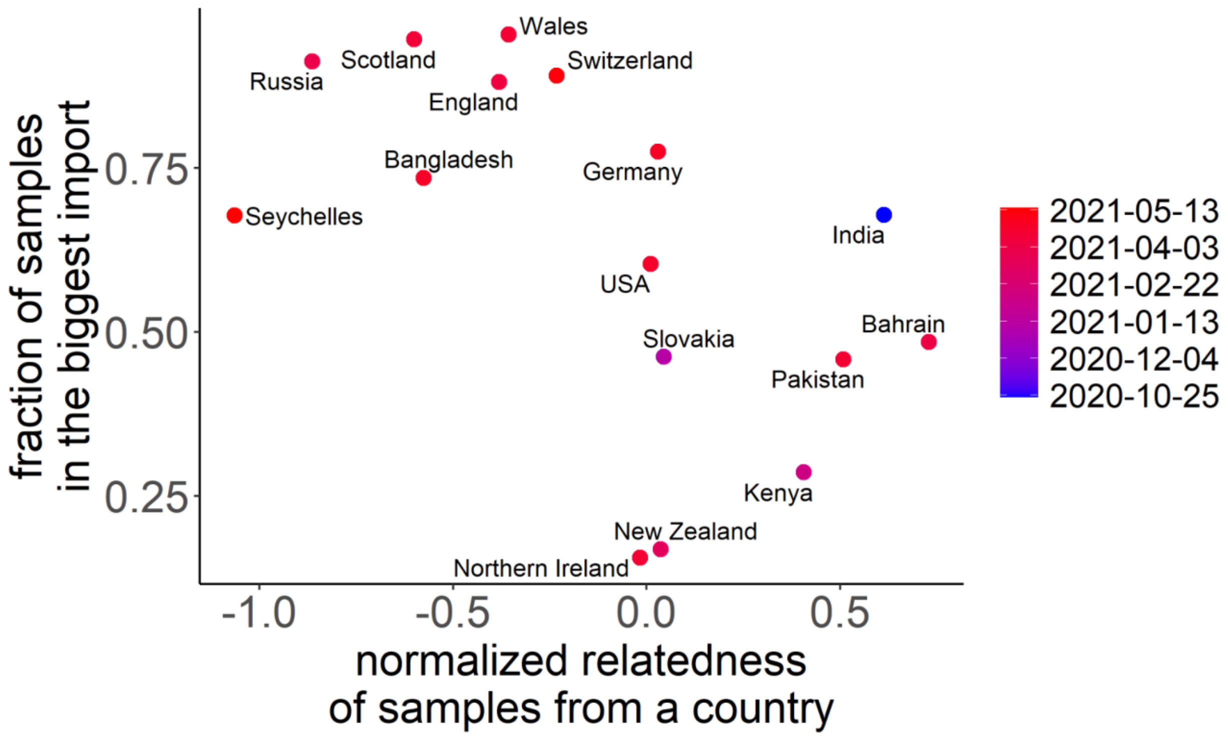

**Figure S10. Russian transmission lineages.** Each horizontal line represents a Russian transmission lineage, ordered by the date of the earliest sample. Circles represent samples taken on a particular date, with circle size representing the number of samples. Circle color indicates the PANGOLIN designation of the corresponding sample.

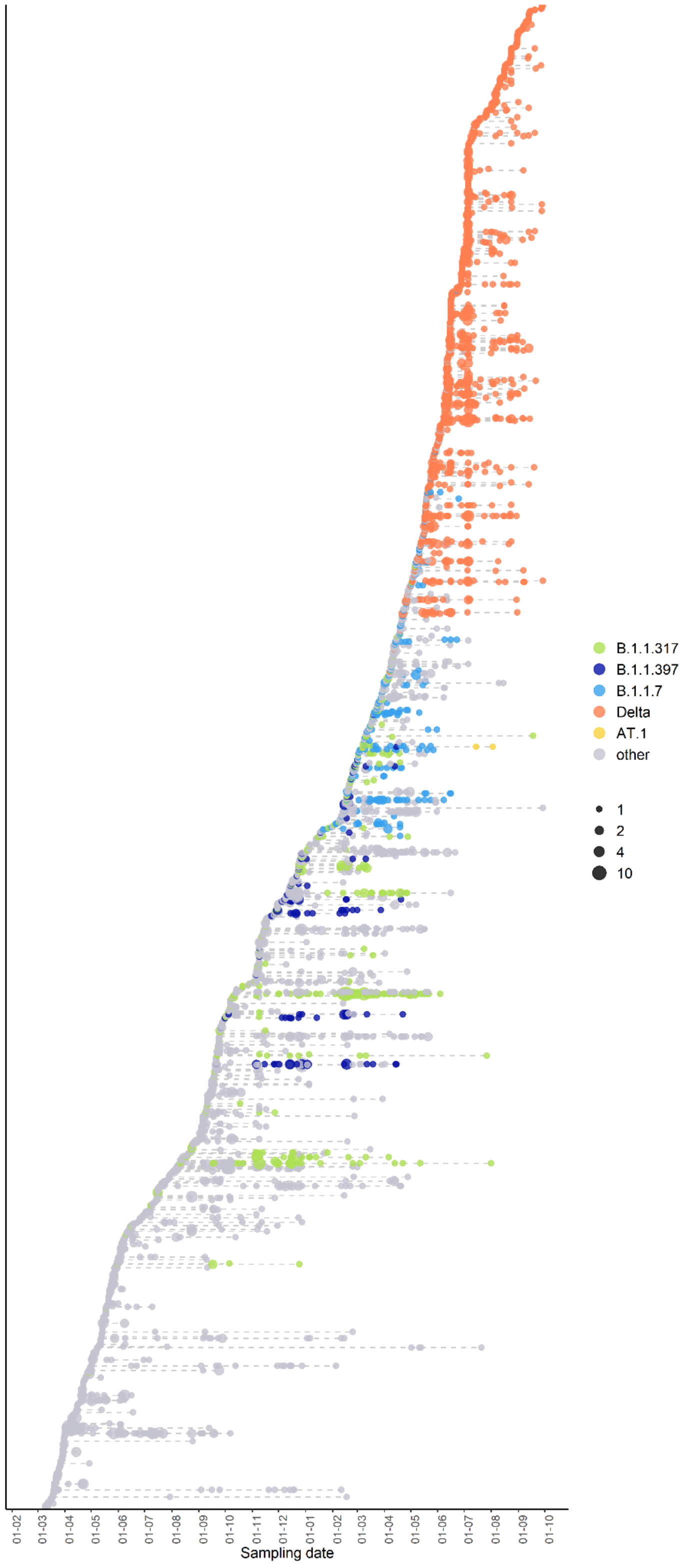

**Supplementary References**


[1] du Plessis L, McCrone JT, Zarebski AE, Hill V, Ruis C, Gutierrez B, et al. Establishment and lineage dynamics of the SARS-CoV-2 epidemic in the UK. Science 2021;371:708–12. https://doi.org/10.1126/science.abf2946.

[2] Stadler T. Phylodynamic Analyses of outbreaks in China, Italy, Washington State (USA), and the Diamond Princess n.d.
