## Supplementary material for "The rise and spread of the SARS-CoV-2 AY.122 lineage in Russia": List of CORGI members

### CoRGI consortium


**Smorodintsev Research Institute of Influenza, WHO-recognized National Influenza Center of Russian Federation (Saint Petersburg, Russia)**

Dmitry Lioznov

Daria Danilenko

Andrey Komissarov

Artem Fadeev

Anna Ivanova

Kseniya Komissarova

Mariia Sergeeva

Maria Pisareva

Veronica Eder

Tamila Musaeva

Maria Timofeeva

Andrey Ksenafontov

Nikita Yolshin

Petr Nekrasov

**Helix LLC (Saint Petersburg, Russia)**

Yury Andreichuk

Dmitry Denisov

Daria Sautkina

**National Research Center for Epidemiology and Microbiology named after Honorary Academician N.F. Gamaleya (Moscow, Russia)**

Alexander Gintsburg

Vladimir Gushchin

Inna Dolzhikova

Denis Logunov

Nadezhda Kuznetsova

Elena Shidlovskaya

Alexey Shchetinin

Maria Nikiforova

Andrei Siniavin

Andrey Pochtovyy

**UMMC-Health LLC (Yekaterinburg, Russia)**

Tatiana Platonova

Mikhail Sklyar

Elena Karbovnichaya

Artur Vorobyov

**Eurasian Association of Therapists**

*Pirogov Russian National Research Medical University (Moscow, Russia)*

Alexander Arutyunov

*Privolzhsky Research Medical University (Nizhny Novgorod, Russia)*

Ekaterina Tarlovskaya

*City Clinical Hospital No. 7, Kazan, Tatarstan*

Zulfia Kim

*Petrozavodsk State University (Petrozavodsk, Russia)*

Tatiana Kuznetsova

*Krasnoyarsk State Medical University (Krasnoyarsk, Russia)*

Marina Petrova

*Privolzhsky Research Medical University (Nizhny Novgorod, Russia)*

Alexandra Vaisberg

### CORGI contributors (in alphabetical order)

**Amur Regional Infectious Diseases Hospital (Blagoveshchensk, Russia)**

Natalia Polovitsa

**Astrakhan State Medical University (Astrakhan, Russia)**

Olga Bashkina

Elena Vakalova

Oleg Rubalsky

Marina Samotrueva

Nikolay Kostenko

**Botkin Memorial Hospital for Infectious Diseases (Saint Petersburg, Russia)**

Denis Gusev

Maria Vashukova

Evgeny Venev

**Centre for Hygiene and Epidemiology in Astrakhan Region (Astrakhan, Russia)**

Elena Vokalova

**Center for Hygiene and Epidemiology in Belgorod Region (Belgorod, Russia)**

Liudmila Berdinskikh

**City Hospital No 40 (Saint Petersburg, Russia)**

Sergey Scherbak

Olga Shneider

Margarita Korzhanova

**Gabrichevsky Moscow Research Institute for Epidemiology and Microbiology (Moscow, Russia)**

Vladimir Aleshkin

Evgenii Rubalsky

**Infectious Diseases Clinical Hospital №1 (Moscow, Russia)**

Svetlana Smetanina

Natalia Antipyat

Marina Bazarova

Aleksandr Shagaev

**Kamchatka Regional Children's Infectious Diseases Hospital (Petropavlovsk-Kamchatsky, Russia)**

Svetlana Shapovalenko

**Murmansk Regional Center for Specialized Medical Care (Murmansk, Russia)**

Elena Labintseva

**Office of the Federal Service for Surveillance on Consumer Rights Protection and Human Wellbeing in the Astrakhan Region (Astrakhan, Russia)**

Natalia Nikeshina

**Ogarev Mordovia State University (Saransk, Russia)**

Irina Stepanenko

**Omsk Research Institute of Natural Focal Infections (Omsk, Russia)**

Valery Yakimenko

Ekaterina Gradoboeva

Aleksei Vasilenko

Elena Poleshchuk

**Orenburg Regional Clinical Infectious Diseases Hospital (Orenburg, Russia)**

Svetlana Soboleva

Denis Durchenkov

Irina Samoryadova

**Peoples' Friendship University of Russia (RUDN University)(Moscow, Russia)**

Olga Burgasova

Valeria Bacalin

Mikhail Odnoralov

Liubov Genralova

**Republican Medical Genetics Center (Ufa, Bashkortostan, Russia)**

Ildar Minniakhmetov

Rita Khusainova

**Saint Petersburg State University (Saint Petersburg, Russia)**

Alexey Masharsky

Maria Baturova

Oula Mansour

**SOGAZ PROFMEDICINA LLC (Saint Petersburg, Russia)**

Galina Razbitskova

Oleg Lezhnev

Larisa Gritsay

**Specialized Clinical infectious Diseases Hospital (Krasnodar, Russia)**

Elena Yakovchuk
