## Supplementary material for "The rise and spread of the SARS-CoV-2 AY.122 lineage in Russia": List of CRIE members

**Federal Budget Institution of Science “Central Research Institute for Epidemiology” of the Federal Service for Supervision of Consumer Rights Protection and Human Welfare (Rospotrebnadzor), Moscow, Russia**

Akimkin V.G.,

Borisova N.I.,

Budkina A.Y.,

Buharina A.Y.,

Bulanenko V.P.,

Cherkashina A.S.,

Chudinov I.K.,

Dolotova S.M.,

Golubeva A.G.,

Goncharov S.E.,

Gulboy E.Y.,

Kaptelova V.V.,

Khafizov K.F.,

Kolesnikov A.A.,

Kondrasheva L.Y.,

Korneenko E.V.,

Kotov I.A.,

Marakulina D.A.,

Mikheeva O.O.,

Mityanina Y.A.,

Nadtoka M.I.,

Noskova O.M,

Roev G.V.,

Saenko S.S.,

Samoilov A.E.,

Selezov S.Y.,

Shipulina O.Y.,

Sinitsyn S.O.,

Smirnova Y.S.,

Solovieva E.D.,

Speranskaya A.S.,

Svetlichnyj D.V.,

Tivanova E.V.,

Valdohina A.V.,

Vyhodceva A.V.,

Zotova M.I.,

Zuev S.N.
